# Time’s up: Using data-driven phenotype-severity metrics not time to map progression in the dementias

**DOI:** 10.64898/2026.01.28.26344700

**Authors:** Viktorija Smith, Rahel Schumacher, Siddharth Ramanan, Arabella Bouzigues, Lucy L. Russell, Phoebe H. Foster, Eve Ferry-Bolder, John C. van Swieten, Lize C. Jiskoot, Harro Seelaar, Raquel Sanchez-Valle, Robert Laforce, Caroline Graff, Daniela Galimberti, Rik Vandenberghe, Alexandre de Mendonça, Giuseppe di Fede, Isabel Santana, Alexander Gerhard, Johannes Levin, Benedetta Nacmias, Markus Otto, Maxime Bertoux, Thibaud Lebouvier, Simon Ducharme, Chris R. Butler, Isabelle Le Ber, Elizabeth Finger, Maria Carmela Tartaglia, Mario Masellis, Matthis Synofzik, Fermin Moreno, Barbara Borroni, Jonathan D. Rohrer, James B. Rowe, Matthew A. Lambon Ralph

**Author notes:** Correspondence to: Matthew Lambon Ralph, Full address: MRC Cognition and Brain Sciences Unit, University of Cambridge, Cambridge, CB2 7EF, UK, Viktorija Smith, Full address: MRC Cognition and Brain Sciences Unit, University of Cambridge, Cambridge, CB2 7EF, UK.

## Abstract

Temporal measures, such as time from diagnosis or symptom onset are often used to track disease severity in neurodegenerative diseases. Due to variations in symptom awareness, clinical presentation timing, diagnostic delays, and disease progression rates, these temporal proxies introduce substantial variance and bias, making it very difficult to map progression clearly and accurately, and to severity-match across contrastive patient groups. To address this challenge, we explored a data-driven approach to derive a transdiagnostic severity metric that is independent of time and, instead, treats temporal metrics as observed, dependent data.

We analysed data from the Genetic Frontotemporal Dementia Initiative (GENFI 1 and 2). We entered neuropsychological scores for symptomatic individuals including any visits prior to conversion from at-risk to symptomatic (n = 265, 522 visits) in an unrotated principal component analysis to derive a transdiagnostic phenotype-severity model. A single component emerged (Kaiser-Meyer-Olkin = 0.92), explaining 65% of the variance, with all neuropsychological assessments loading highly. This global severity component fitted the data equally well across genetically or clinically defined groups, as well as severity levels.

The severity measure’s validity was supported by a clear relationship with the Clinical Dementia Rating scale, and its stability was confirmed when a much broader range of neuropsychological and behavioural measures were included. Additionally, the severity score accounted for a high portion of the total variance in neuropsychological test scores, substantially more than the low proportion accounted for by standard temporal measures. To derive a time-efficient sub-battery, we demonstrated that three neuropsychological assessments (Digit Symbol, Verbal fluency (letters) and Trail Making Test- Part B were able to explain the majority of unique variance in cognitive severity. Finally, by treating time as an observed dependent variable, we showed that the baseline velocity (change in severity measure over time) varied by genetic group, with progranulin mutation carriers being the fastest.

This data-driven approach provides an objective, precise measure of disease severity and progression, and it may shed new light on when clinical heterogeneity reflects distinct subtypes rather than differences in disease stage.

## Introduction

Sensitive measures that span the full range of severity in neurodegenerative diseases are required to aid diagnosis, therapeutic management and development of effective clinical trials.^1–3^ Commonly used temporal markers, such as time from diagnosis or symptom onset, are plagued with multiple challenges relating to variability in patients’/carers’ symptom awareness, timing of clinical presentation, diagnostic delays, and rates of disease progression.^4–6^ Biomarkers may exacerbate the challenge, rather than resolve it, if they have differential sensitivity and rates of emergence during disease progression.^7^ As a result, temporal proxies of severity reduce the ability to align individuals at the same stage of the disease, which makes it difficult to separate qualitatively different subtypes of patients from quantitatively varying severity levels of a single disease trajectory.^8,9^ Precise measures of severity or disease stage would improve clinical trials through better control of severity-related effects within and across subtypes of patients, as well as providing outcome measures that are centred on the core nature of the disease.^3^ Moreover, instead of using time as the severity proxy, a non-temporal severity measure would allow time to become an observed “dependent” variable and would allow one to compare rates of progression across clinical subtypes. Accordingly, the core goal of this study was to evaluate a novel approach that uses individuals’ performance on detailed neuropsychological measures to develop a data-driven transdiagnostic metric of clinical severity, which is not dependent on temporal progression.

Research studies often rely on time-based or coarse scales of independent functioning to help map pathophysiological processes or control for severity-related effects on underlying mechanisms.^10–12^ Given that neurodegenerative processes are intrinsically linked to passing time, it seems intuitive to use actual or estimated time as a proxy for disease severity. However, if temporal measures are used as proxies for disease severity, this introduces significant limitations. Unlike acute onset neurological disorders (e.g., stroke or traumatic brain injury events), it is difficult to pinpoint precise dates for disease or symptom onset, as the biological changes underlying the disease are gradual. Temporal measures introduce broad heterogeneity as patients, families and clinicians are differentially aware of symptoms and their relevance to subsequent diagnosis. The sensitivity to clinical changes and seeking of medical advice varies between patients and is linked to education, marital status, knowledge and emotional attitude.^4,13^ The referral pathway and entry to medical services and clinical trials also varies; therefore, the moment of entry is not directly comparable across individuals. In addition, clinical progression does not follow a uniform monotonic pattern and individuals progress at variable rates.^2,14^ Due to these challenges, effective and precise methods are still required to match individuals based on their disease severity level. The ability to measure disease progression precisely has implications for research (i) to understand the natural history of symptom onsets at different levels of severity within the same disease and (ii) to match contrastive groups with a generalised metric of severity to improve differentiation of phenotypes and subtypes of disease. Recruitment into clinical trials also relies on the ability to stratify patients according to severity with the aim to reduce clinical heterogeneity amongst groups. Precise measures of disease progression are key in clinical trial outcomes, especially where a disease modifying therapy has an optimal window in the disease course. The identification of measures that are able to capture subtle cognitive changes before clinical diagnosis of dementia has also been key due to the focus on early interventions.^15–17^

Here, we propose an alternative data-driven approach using principal component analysis (PCA) to derive a transdiagnostic phenotype-severity metric.^18^ We test this approach in patients with or at risk of frontotemporal dementia (FTD) from the Genetic Frontotemporal Dementia Initiative (GENFI) study, due to autosomal dominant mutations in C9orf72, progranulin (GRN), and microtubule-associated protein tau (MAPT) genes.^19^ Frontotemporal dementias are a group of heterogeneous disorders characterized by neurodegeneration including the frontal and temporal lobes with a spectrum of changes in behaviour, language and motor function. The GENFI cohort is ideally suited to evaluate the transdiagnostic phenotype-severity metric because: (i) it contains detailed formal neuropsychology assessment that can be utilised to develop the data-driven severity metric; (ii) there is a spectrum of subtypes and variations of phenotype in FTD including three genetic subtypes; and (iii) patients are repeatedly tested over time thus generating a rich cross-sectional-longitudinal database for analysis.

Specifically, we determined the extent to which a data-driven PCA severity model explains variance in clinical scores in carriers of C9orf72, GRN, or MAPT mutations, and across diagnoses of behavioural variant FTD (bvFTD), primary progressive aphasia (PPA), and amyotrophic lateral sclerosis (ALS). We compare the PCA-derived measure to standard temporal markers and clinical rating scales to estimate the rates of progression by gene and phenotype, and then identified which subset of neuropsychological tests are most time efficient for the detection of early clinical changes.

## Methods

### Participants

We examined GENFI data collected between January 2012 and February 2021. Across the GENFI 1 and GENFI 2 waves, data from 1130 unique participants (asymptomatic and symptomatic) totalling 2861 visits were available. We focused on GENFI participants who were symptomatic carriers of a common pathogenic mutations (C9orf72, GRN or MAPT) or first-degree relatives of a known symptomatic carrier.^19^ Symptomatic status is based on standardised clinical assessment consisting of a medical history, family history, and physical examination. To capture longitudinal changes and to maximise sampling across the full phenotypical severity spectrum, we included patients who had at least three complete neuropsychological assessments. The resultant dataset comprised 1678 visits from 686 individuals (Table 1). Given their small sample size, participants with a clinical diagnosis of Parkinson’s Disease, Mild Cognitive Impairment, Alzheimer’s Disease, Progressive Supranuclear Palsy, Corticobasal Syndrome, Bipolar Disorder and non-specified dementia were combined into the ‘Other’ category.

**Table 1:** Demographics and clinical scores at baseline across at-risk and symptomatic groups.

|  | At-risk<br>*n* = 421 | Symptomatic and<br>converters<br>*n* = 265 | Group differences at baseline visit |
| --- | --- | --- | --- |
| Age, mean (SD) | 43.2 (11.4) | 61.7 (8.61) | $t(662.61) = -23.98, P < 0.001$ |
| Sex |  |  |  |
| Male | 167 (39.7%) | 156 (58.9%) | $\chi^2(1) = 23.3, P < 0.001$ |
| Female | 254 (60.3%) | 109 (41.1%) |  |
| Education, mean (SD) | 14.5 (3.34) | 12.4 (3.66) | $t(522.91) = 7.61, P < 0.001$ |
| Genetic group |  |  |  |
| C9orf72 | 172 (40.9%) | 137 (51.7%) | $\chi^2(2) = 8.3, P = 0.012$ |
| MAPT | 72 (17.1%) | 42 (15.8%) |  |
| GRN | 177 (42.0%) | 86 (32.5%) |  |
| CDR®-FTLD |  |  |  |
| 0 | 228 (54.2%) | 12 (4.5%) | $\chi^2(4) = 319.16, P < 0.001$ |
| 0.5 | 62 (14.7%) | 26 (9.8%) |  |
| 1 | 7 (1.7%) | 43 (16.2%) |  |
| 2 | 1 (0.2%) | 50 (18.9%) |  |
| 3 | - | 47 (17.7%) |  |
| Missing | 123 (29.2%) | 87 (32.8%) |  |
| Number of visits, mean (SD) | 2.75 (1.75) | 1.97 (1.40) | $t(522.91) = 7.61, P < 0.001$ |
| Years from onset, mean (SD) | - | 4.43 (3.70) | - |
| Diagnosis |  |  |  |
| bvFTD | - | 160 (60.4%) | - |
| PPA |  | 38 (14.3%) |  |
| ALS |  | 25 (9.4%) |  |
| Other |  | 15 (5.7%) |  |
| MMSE, mean (SD) | 29.3 (1.15) | 23.2 (6.35) | $t(256.98) = 14.89, P < 0.001$ |
| Digit Span Forward, mean (SD) | 7.98 (2.16) | 5.50 (2.46) | $t(508.76) = 13.47, P < 0.001$ |
| Digit Span Backward, mean (SD) | 6.73 (2.18) | 3.73 (2.31) | $t(536.88) = 16.97, P < 0.001$ |
| Digit Symbol, mean (SD) | 56.4 (12.4) | 26.1 (15.1) | $t(481.65) = 27.37, P < 0.001$ |
| Boston Naming, mean (SD) | 27.6 (2.60) | 20.2 (7.77) | $t(301.52) = 15.03, P < 0.001$ |
| Block Design, mean (SD) | 47.2 (14.8) | 18.4 (16.3) | $t(519.8) = 23.73, P < 0.001$ |
| Verbal Fluency - Animals, mean (SD) | 23.9 (5.63) | 11.8 (7.04) | $t(471.61) = 23.61, P < 0.001$ |
| Verbal Fluency - Letters, mean (SD) | 39.5 (12.9) | 18.1 (13.9) | $t(529.27) = 20.19, P < 0.001$ |
| Trail Making – A time, mean (SD) | 28.3 (11.2) | 68.8 (41.8) | $t(287.85) = 15.42, P < 0.001$ |
| Trail Making – B time, mean (SD) | 67.6 (35.1) | 208 (92.3) | $t(312.62) = 23.73, P < 0.001$ |

CDR®-FTLD = Clinical Dementia Rating Scale with the National Alzheimer Coordinating Centre frontotemporal lobar degeneration module; bvFTD = behavioural-variant frontotemporal dementia; PPA = primary progressive aphasia; ALS = amyotrophic lateral sclerosis; MMSE = Mini-Mental State Examination.

Institutional review board approval and informed consent was obtained from all participants at each of the GENFI research sites.

### Assessments

Participants underwent the standardised GENFI clinical assessment including the Mini-Mental State Examination (MMSE), Cambridge Behavioural Inventory-Revised (CBI-R), the Frontotemporal Dementia Rating Scale (FRS) and the Clinical Dementia Rating Scale with the National Alzheimer Coordinating Centre FTLD score (CDR®-FTLD).

Neuropsychological assessments in GENFI 1 and GENFI 2 waves overlap with the Uniform Data Set^20^: Digit Span forwards and backwards from the Wechsler Memory Scale-Revised, a Digit Symbol Task, Parts A and B of the Trail Making Test, the short version of the Boston Naming Test, and Category Fluency (Animals). Additional tests included in the battery were Letter Fluency (‘FAS’) and the Wechsler Abbreviated Scale of Intelligence Block Design^19^.

The GENFI 2 wave also included the Camel and Cactus Test, Color-Word Interference Test from Delis-Kaplan Executive Function System (D-KEFS), the Benson Complex Figure test, the Free and Cued Selective Reminding test, the Faux Pas recognition test, Revised Self-Monitoring Scale (RSMS), modified Interpersonal Reactivity Index (mIRI), and the Facial Emotion Recognition test (FER). These were included together with the initial core neuropsychological battery in a supplementary analysis.

### Data analysis

Statistical analyses were conducted in R Studio 2024.04.2+764. The first aim was to generate a data-driven model of severity. Missing neuropsychological data were imputed using a PCA-based method. A k-fold cross-validated approach was first used to identify the number of dimensions that minimised the mean squared error of prediction when reconstructing the incomplete dataset (Supplementary Table 1). Once the optimal number of components was selected, missing values are iteratively estimated and the PCA recomputed until convergence (or maximum 1000 iterations). We then applied unrotated PCA to the combined all longitudinal GENFI 1 and GENFI 2 data from symptomatic carriers including any visits prior to conversion from at-risk to symptomatic, if applicable. The component explaining the highest variance with an eigenvalue > 1 was extracted and individual participant’s values for this component were used as the measure of global severity for subsequent analysis. To assess the impact of the number or selection of data points, we compared unrotated PCA analysis results when using all data points, only the baseline visits (i.e., cross-sectional only), or a subset of data containing only the first, middle and last visits for each individual (which confirmed that the severity PCA metric was highly stable, see Supplementary Table 2). Using the weightings on each test, we then calculated the severity score for all asymptomatic carriers (scaled test score * test weighting) to compare variation across the severity range. To explore differences in severity score across clinical groups (i.e. symptomatic vs asymptomatic carriers) we performed a t-test and a Welch’s and one-way ANOVA with post hoc Bonferroni and Tukey HSD tests. In order to test the validity of the severity score against an independent measure, we explored its relationship with a functional measure of impairment (CDR®-FTLD). We fitted ordinal regression models to predict CDR®-FTLD scores by the severity score alone or by the severity score plus CBI-R Total score and compared the fit of these models using a likelihood ratio test. To assess how temporal measures related to severity, we applied both frequentist and Bayesian linear regression models to predict severity score by years from symptom onset and visit number.

Next, we used the severity score as the data-organising variable and re-introduced time as an observed measure. To explore the rate of change in severity, we performed 1) quadratic linear regression model with change in severity; and 2) linear mixed effects models with the rate of change (change in severity/time between visits) predicted by the severity score.

To estimate the duration of progression across genetic and diagnostic groups, we applied a linear mixed effects model predicting the rate of change in severity per year as a function of mean severity with genetic group and age at visit as covariates. To calculate the time taken to traverse the severity spectrum, we used the model coefficients for each genetic group to predict the rate of change at 0.1 increments across the severity spectrum, which allowed a calculation of the time taken to progress the severity continuum (time=0.1/rate of change). Low severity scores can fluctuate reflecting normal test-retest variability, therefore to avoid division by zero or negative values, speeds ≤ 0 were excluded, as they reflected improvements in severity. The cumulative time at each severity level for each genetic group was calculated by summing all time increments. This allowed to generate severity-time curves for each genetic group.

To identify sensitive neuropsychological assessments that detect early cognitive changes we looked at the relationships between the raw test scores and the severity factor. By taking the scores at 1.5 SD below the mean for a 72 year old with 12 years of education based on published cut-off scores for each test, we were able to calculate a severity score indicating a level of clinical impairment.^21–27^ This calculated severity score cut-off is a stringent threshold, as for a diagnosis of cognitive impairment it is not necessary to score below 1.5 SD on all administered tests. For each test we identified how individual scores fall above or beyond this cut-off point in comparison to the test specific cut-off scores (1.5 SD below the mean). McNemar test was applied to compare differences between two score quadrants for all tests: 1) above the impaired threshold for test, but high severity score 2) below the impaired threshold for test, but low severity score. This assessed the difference in the test detection rate of false negatives and false positives, with the latter indicating cases where individuals are scoring poorly on the test despite overall global severity still intact.

The above analyses were performed on the shared neuropsychological battery combined across GENFI 1 and GENFI 2. To explore the influence of additional behavioural and functional measures, we explored the effect of including CBI-R and FRS in our initial PCA model and performed another unrotated PCA on the GENFI 2 dataset only.

### Data availability

Anonymised data can be shared according to the GENFI data sharing agreement, after review by the GENFI data access committee with final approval granted by the GENFI steering committee. To apply, please see https://www.genfi.org/.

## Results

### Demographics

Compared to the at-risk participants, symptomatic and converted individuals were on average older, more likely to be male, and had fewer years of education (Table 1). Diagnosis of the behavioural variant of frontotemporal dementia was the most common (60.4%). As expected, the groups differed significantly across all functional and neuropsychological measures (Table 1).

### Capturing global severity

For our first aim, we tested whether a data-driven PCA severity model can explain the majority of variance in the neuropsychological scores. When the neuropsychological assessment data from GENFI 1 and GENFI 2 were entered into an unrotated PCA, all tests loaded highly on the first factor (Kaiser-Meyer-Olkin = 0.92, Supplementary Table 2). As the only factor with an eigenvalue > 1 and explaining 65% of variance, it can be interpreted as reflecting each individual’s overall cognitive severity. All neuropsychological tests had a high loading >= 0.78 except for Digit Span Forward and Boston Naming Test which were still high ∼0.6. There was very little variation in the PCA outcome whether using all data, first visit only, or first, middle and last visits only (see Supplementary Table 2), therefore we completed all subsequent analysis using all the available data.

As expected, the symptomatic group had a significantly and considerably higher severity score compared to the at-risk individuals (*t*(604.6) = -39.30; *P* < 0.001, Figure 1A). The severity score was significantly related to the CDR®-FTLD score (*β* = 1.723, *SE* = 0.075, *z* = 22.97, *P* < .001, *AIC* = 2308.97; McFadden’s pseudo *R*^2^ = 0.243 (Figure 1B)). For every one unit increase in severity score, the odds of having a higher CDR®-FTLD score increased by a factor of 5.60 (95% CI: [4.84, 6.49]). Note that a broad range of severity scores was associated with each CDR®-FTLD score from none to severe impairment, suggesting that the clinical ratings (CDR scale) capture challenges beyond neuropsychological severity alone (Figure 1). This was confirmed by examining the Cambridge Behavioural Inventory-Revised scores, which also showed a strong association with CDR®-FTLD score (*β* = 0.087, *SE* = 0.004, *z* = 24.12, *AIC* = 1953.90; McFadden’s pseudo *R^2^* = 0.360). For every one unit increase in severity score, the odds of having a higher CDR®-FTLD score increase by a factor of 1.09 (95% CI: [1.08, 1.10]). The combined model with CBI-R and neuropsychological severity variables showed improved fit, with both severity score (*β* = 0.866, *SE* = 0.088, *z* = 9.86) and CBI-R (*β* = 0.071, *SE* = 0.004, *z* = 18.23) remaining statistically significant (*AIC* = 1854.70, McFadden’s pseudo *R^2^* = 0.393). The model with both predictors significantly outperformed the neuropsychological severity score only model, suggesting that clinicians’ CDR®-FTLD ratings are, as expected, influenced by both neuropsychological severity and behavioural changes (*χ²*(1) = 456.27, *P* < 0.001).

**Figure 1:**
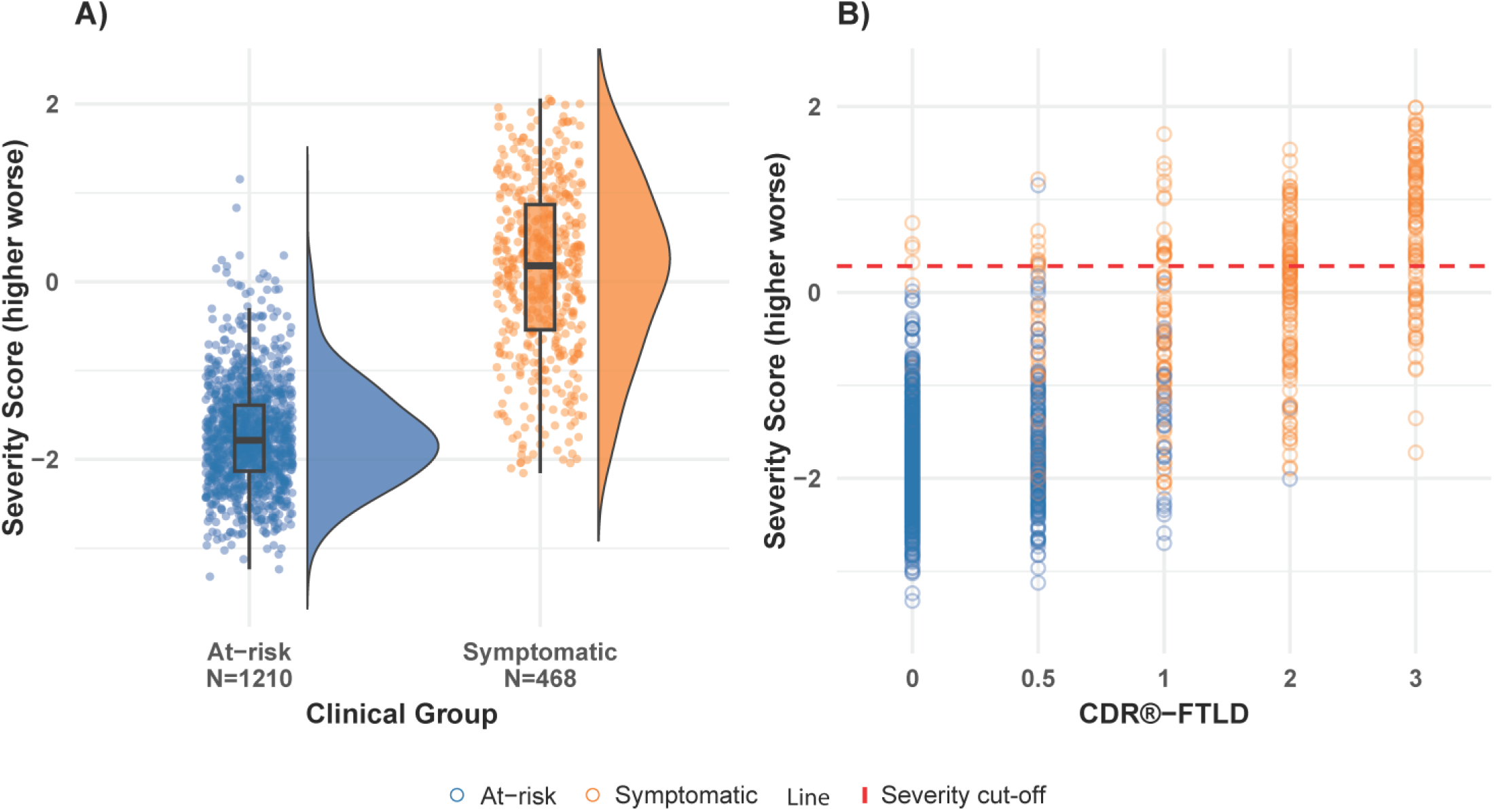
Severity score associations by clinical group (A) and functional impairment (B). **(A)** Distribution of severity scores among all visits for at-risk (blue) and symptomatic (orange) individuals. **(B)** The relationship between severity scores and CDR®-FTLD scores for at-risk (blue) and symptomatic (orange) individuals. The red dashed line represents the severity score cut-off indicating a level of clinical impairment.

Within symptomatic individuals, the measure captured the full range of severity for all genetic and diagnostic groups (Figure 2). Significant differences in severity score were observed across genetic groups (*F*(2,225.16) = 16.09, *P* < 0.001) with MAPT (*m* = -0.50, *SD*=0.93) lowest compared to GRN (*m* = 0.17, *SD* = 1.14) and C9orf72 (*m* = 0.08, *SD* = 0.88). Similarly, the PPA diagnostic group (*m* = 0.68, *SD* = 0.97) was significantly worse than all other groups (*F*(3,464) = 9.207, *P* < 0.001): bvFTD (*m* = 0.07, *SD* = 0.95), ALS (*m* = -0.13, *SD* = 0.76) and ‘Other’ (*m* = -0.07, *SD* = 0.93, Figure 2).

**Figure 2:**
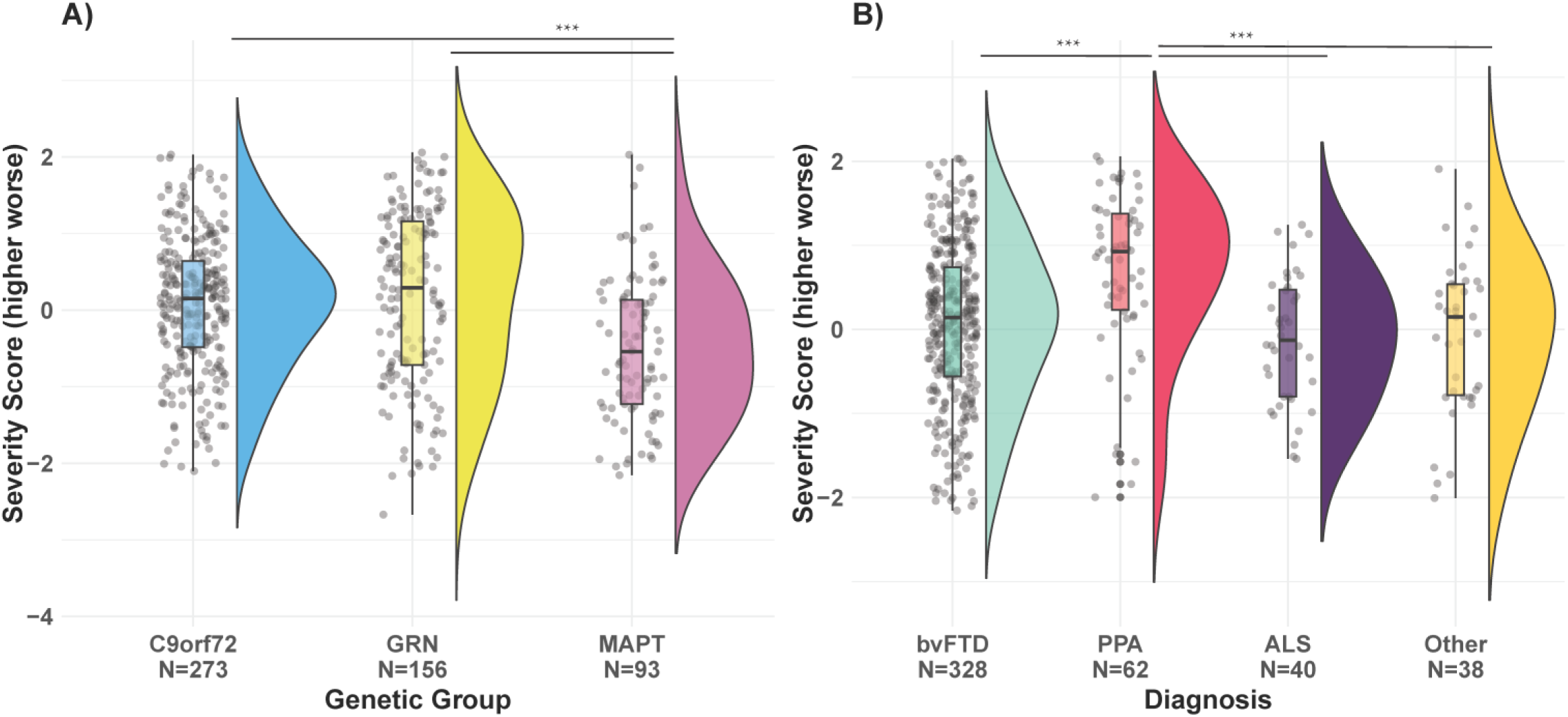
Distribution of severity scores for symptomatic individuals across genetic (A) and diagnostic groups (B). C9orf72 - Chromosome 9 open reading frame 72, GRN – progranulin, MAPT - Microtubule-Associated Protein Tau, bvFTD - behavioural variant frontotemporal dementia, PPA – primary progressive aphasia, ALS - amyotrophic lateral sclerosis. *Note:* *** P < 0.001.

### Severity and its relationship to temporal measures

We compared the data-driven severity measure to commonly-used temporal measures. First, we examined years since reported symptom onset. Despite spanning the full range of severity scores across all three genetic subgroups, there was almost no relationship with the reported time since onset with moderate Bayes factor evidence for the absence of such a correlation (*BF* = 0.261, *β* = -0.013; *t*(466)= -1.387, *P* = 0.166, Figure 3A). Indeed, the *R²* value was very small (*R²* <= 0.005) with only 0.41% of the variance in severity score explained by years from onset. Within the bvFTD group with C9orf72 expansions, a subgroup of individuals retained a low severity score even after over 20 years since diagnosis, suggesting a slowly-progressive neuropsychiatric and non-demented subgroup.

**Figure 3:**
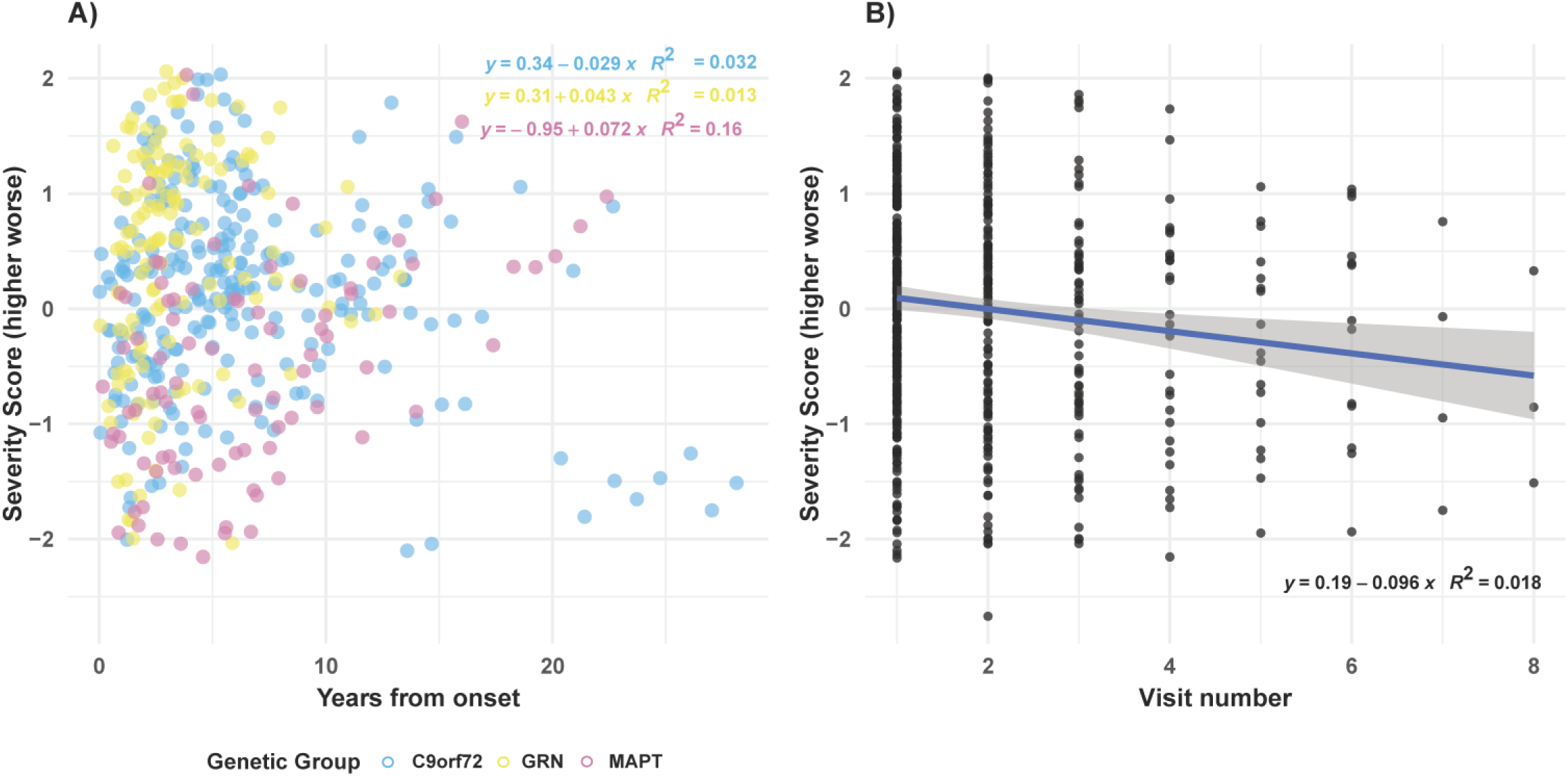
Severity score associations with years from symptom onset (A) and visit number (B). **(A)** The relationship between severity score (y-axis) and years from symptom onset (x-axis) in symptomatic carriers and converters, stratified by genetic group. Linear regression models were fit separately for each genetic group. The equations and *R²* values show the estimated slopes and model fit for each group’s severity trajectory over time. **(B)** Severity score plotted against visit number in symptomatic carriers and converters. A linear model was fitted across all visits. The shaded area represents the 95% confidence interval around the fitted line.

When the data were grouped by a different, temporal measure, ‘visit number’ that is commonly used in longitudinal cohort studies, a significant impact of attrition bias was also observed (Figure 3B). At the time of initial recruitment to GENFI, the patients spanned the full range of severity; counterintuitively, the mean severity score then reduced over testing rounds. This was largely due to patient attrition, which occurred across the full range of severity but was more likely for the severe cases.

### Rates of progression across the severity space

First, we evaluated the relationship between degree of change (the difference between consecutive pairs of severity scores) and the patients’ level of severity. Across all at-risk and symptomatic carriers with longitudinal data the overall model was statistically significant (*F*(2,989) = 125.8, *P* < 0.001) and accounted for 20.1% of variance in severity change. A significant positive quadratic effect was observed (*β* = 0.04, *SE* = 0.01, *t* = 4.75, *P* < 0.001) reflecting that the degree of severity change increased with severity (Figure 4).

**Figure 4:**
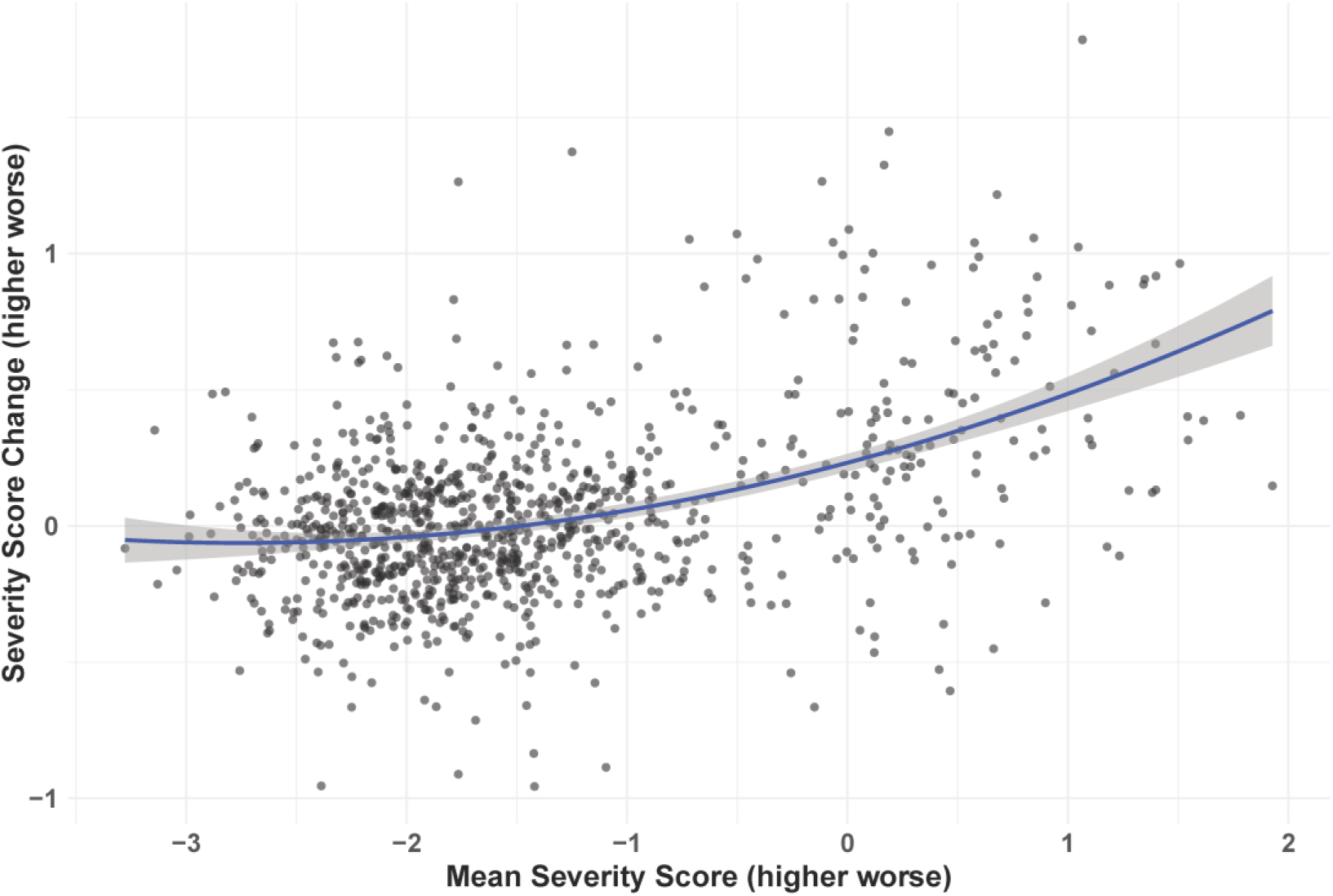
Severity score change along the continuum. A second-order quadratic regression model was used to investigate the non-linear relationship between severity score change over consecutive visits along the severity continuum. The fitted curve is shown in blue with 95% confidence interval shaded grey. The model showed a significant quadratic effect with more change occurring as severity increased.

Secondly, by dividing the severity score change by years between visits, we explored the patients’ “speed” across the severity spectrum, separately for each genetic group (Figure 5). Progression was found to accelerate as individuals became severer. The GRN genetic group had a higher speed on entry (higher intercept) compared to the C9orf72 mutations carriers (*β* = -0.236, *t*(91.38) = -4.01, *P* < 0.001). When controlling for age, this remained significant between GRN and C9orf72 genetic groups (*β* = -0.242, *t*(92.94) = -4.09, *P* < 0.001), however MAPT mutation carriers were also significantly lower compared to GRN (*β* = -0.176, *t*(75.25)= -2.19, *P* < 0.04). This difference probably emerged due to the lower mean age of MAPT group (*m* = 56.6) compared to GRN mutations carriers (*m* = 64.8).

**Figure 5:**
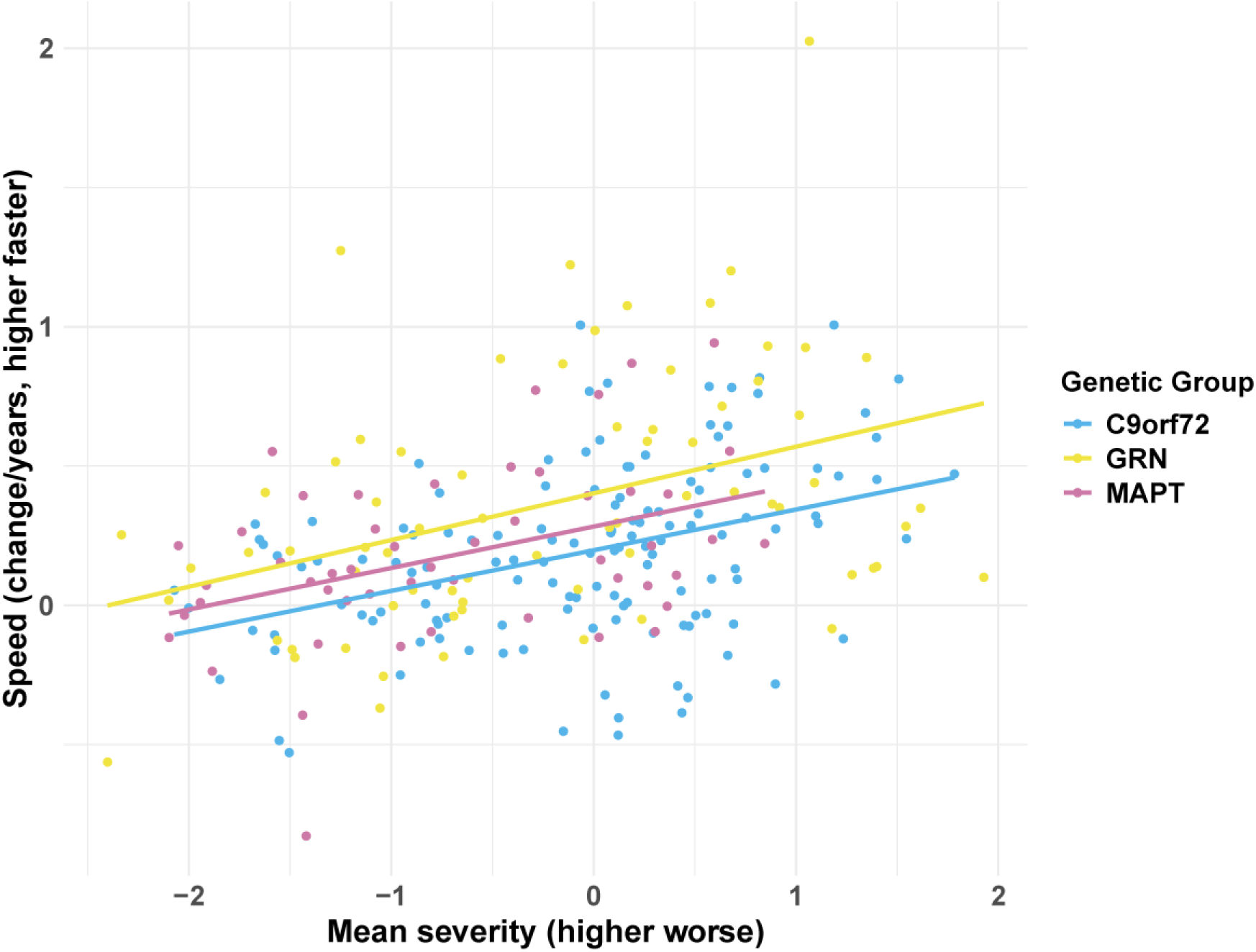
The "speed" of change, defined as the severity change between consecutive visits over time, association with the mean severity of those visits. After controlling for participant random effects and genetic group and age as covariates, a significant positive association emerged, indicating that progression accelerates as impairment increases across genetic groups.

Using these observed “speeds” at each level of severity, it was possible to estimate the time taken to transverse across the severity spectrum for each genetic group (Figure 6). GRN mutation carriers were estimated to take 6.8 years to transverse the severity space from -1.2 to 2, whereas C9orf72 and MAPT groups were estimated to take 19.7 and 11.9 years respectively. As can be observed in Figure 6, these nonlinear functions imply that people spend a considerable amount of time in the “normal-to-prodrome” ranges of severity and less time in the “symptomatic” range. In order to estimate the time of symptomatic onset, Figure 6 shows a projection from the normative control cut-off severity score (0.142) to cumulative years for each group (GRN = 3.8yrs; MAPT = 7.7yrs; C9orf72 = 14.6yrs). Accordingly, the time that each genetic group spends in the symptomatic range (i.e., estimated by the time taken to reach a severity score of 2) was much closer across the groups (GRN = 3.0yrs; MAPT = 4.2yrs; C9orf72 = 5.1yrs).

**Figure 6:**
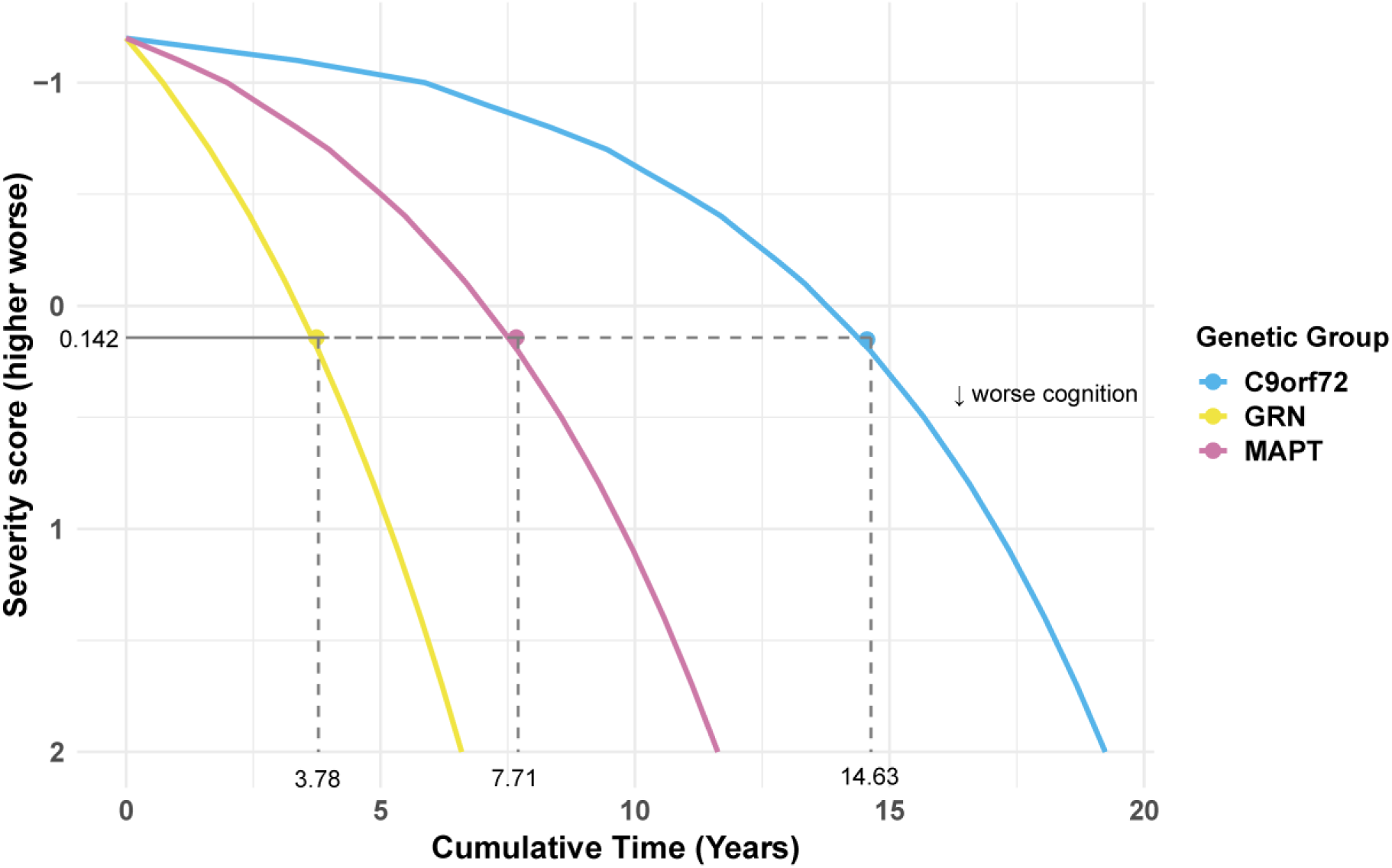
Estimated severity progression plots show the cumulative time required for progression along the severity continuum by genetic group. Each line depicts model-derived severity trajectories over time, based on the predicted progression rate across severity intervals within each genetic group. The y-axis depicts severity, with higher values indicating more impairment. The axis is reversed to reflect increasing severity downward. The x-axis depicts the total amount of time (in years) required to move from least to most severe on the severity continuum. The dashed lines project from the normative severity score cut-off (*m* = 0.142, mean) to the time point when this threshold is reached for each genetic group.

### Test selection for efficient assessment and sensitivity to early clinical changes

In longitudinal studies and clinical trials, time for neuropsychological assessments is limited thus requiring a careful selection of tests. The individual tests’ loadings on the severity score are not a useful selection criterion as all tests loaded similarly highly. Therefore with a view to a time-efficient yet accurate reduced GENFI battery, we explored which of the fastest tests explained the most unique variance. Stepwise forward regression models with severity score as the outcome, showed that a combination of three time-efficient cognitive tests together explained ∼93% of the variance. First, Digit Symbol alone accounted for the majority of variance (*R²* = 0.81, *F*(1, 520) = 2227, *P* < 0.001). Next, adding Verbal fluency (letters) explained an additional 9% (*R²* = 0.895, *F*(2, 519) = 2215, *P* < 0.001), with Trail Making Test – Part B accounting for another 3% (*R²* = 0.931, *F*(3, 518) = 2354, *P* < 0.001). All other tests explained ≤ 2% of the remaining variance. These three tests take around 4 minutes each to complete, and thus could be an efficient core battery for capturing cognitive severity.

With the aim of identifying measures sensitive for detecting early clinical changes, we explored the relationship of the severity score with the raw scores on the individual neuropsychological tests (Figure 7) (see Supplementary Figure 1 and Supplementary Figure 2 for subgrouping according to genetic status and diagnostic group).

**Figure 7:**
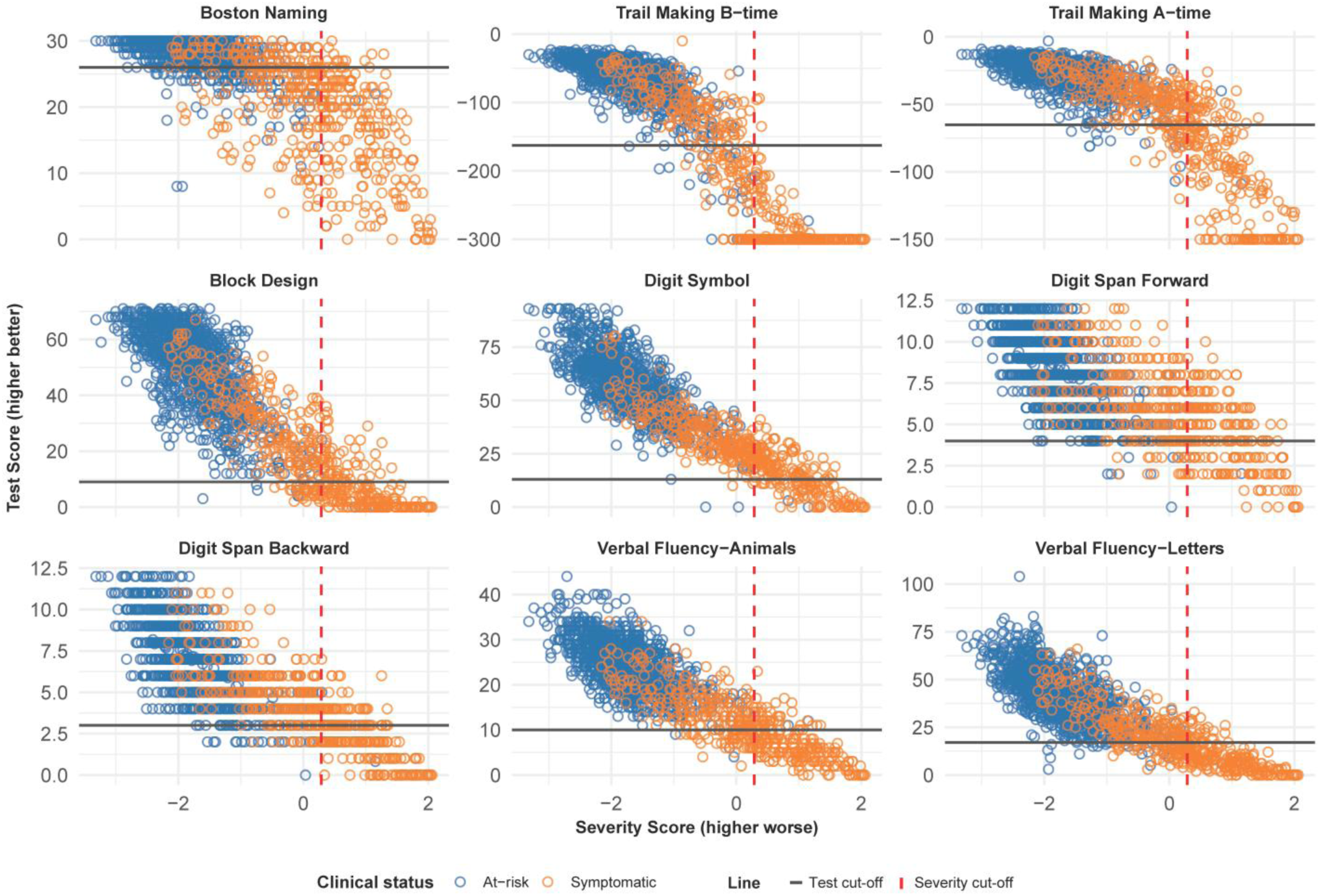
Association of the overall severity score with raw scores of neuropsychological tests and their cut-offs. Each scatterplot includes data across all visits for symptomatic (orange) and at-risk (blue) individuals. A lower severity score indicates less impairment. Black horizontal lines represent the test score cut-off selected at 1.5 SD below the mean for a 72 year old with 12 years of education. The vertical red dashed line is the severity cut-off calculated by all test score cut-off’s multiplied by test weightings on the first component.

The Trail Making Test – Part B may be a potentially useful test to detect early decline, as patients low in overall severity score were already scoring below the control range for this test (i.e., they fall into the lower-left quadrant of the figure; *χ²*(1, 1678) = 109.4, *P*< 0.001, see Supplementary Table 3). This included a subset of asymptomatic individuals, who were noted as symptomatic at subsequent visits. The Boston Naming Test also showed a similar pattern but this was largely due to the MAPT genetic carriers who are known to have early anomia (see Supplementary Table 3).^28,29^

### Effects of extending the neuropsychological battery

To test the stability of the PCA-computed score, we explored what happens to the computed severity score when the core neuropsychology battery is augmented in two different ways. First, we examined what happens if a greater number and wider variety of neuropsychological tests are included. During the second wave of the GENFI study, the neuropsychological battery was extended. The PCA severity score from this extended GENFI2 battery was similar to the initial GENFI-1 PCA, with again most tests loading highly on the first factor (explaining 53% of the variance; Kaiser-Meyer-Olkin=0.94, see also Supplementary Table 4). The Modified Interpersonal Reactivity Index and the Revised Self-Monitoring Scale, two questionnaires completed by carers had lower loadings at 0.416-0.537. As a subset of individuals completed both the initial GENFI 1 and extended GENFI 2 battery (*n* = 200, 370 visits), we were able to compare the two derived severity scores across the same individuals. The severity scores from the two batteries were extremely highly correlated, reflecting that augmenting the battery had limited impact on the severity score (see Supplementary Figure 3A).

Second, FTD leads to behavioural changes as well as cognitive changes. This may explain why some individuals had low severity scores but high CDR®-FTLD scores. To test the effects of behavioural measures on the severity score we ran another PCA including CBI-R and FRS measures. The first component was dominant and explained 58% of variance (Kaiser-Meyer-Olkin=0.90, details in Supplementary Table 5). All tests (including behavioural CBI-R and FRS) loaded highly onto it. A weaker second component (14%) emerged with highest loadings on CBI-R (0.736) and FRS (0.7) (i.e., additional variance in behavioural scores). Both this first “generalised severity” component (*β* = 1.437, *SE* = 0.117, *z* = 12.31, *P* < .001, *AIC* = 1013.3; McFadden’s pseudo *R^2^* = 0.155) and the second “behavioural change” component (*β* = 0.983, *SE* = 0.108, *z* = 9.11, *P* < .001, *AIC* = 1104.9; McFadden’s pseudo R^2^ = 0.078) were significant predictors of the CDR®-FTLD score. The combined model with both components showed improved fit (*AIC* = 868.92, McFadden’s pseudo *R^2^* = 0.278). This further supports the previous results, that CDR®-FTLD captures both cognitive and behavioural severity. The relationship between the first PCA severity scores derived from cognitive scores only or with CBI-R and FRS added, remained extremely correlated, with a slope of 0.987 and an *R^2^* of 0.97 (Supplementary Figure 3B). This strong relationship suggests that the first component continues to reflect the overall severity dimension.

## Discussion

This study introduces a solution to the poor reliability and interpretability of standard temporal measures, such as time since diagnosis or symptom onset. Time-based metrics are a poor reflection individual patients’ symptom severity and progression. Instead, we present a data-driven, transdiagnostic phenotype-severity metric, illustrated in the context of genetic FTD. This measure offers a direct representation of disease state, capturing variation in cognitive and behavioural features across individuals. Such a data-driven approach to quantify severity offers several advantages: i) the measure is not dependent or restricted by the limitations of time; ii) it is independent of diagnosis or other underlying clinical features (e.g., genetics); iii) it is objective and can combine multiple test scores for increased reliability. All these features allow us to place an individual’s profile on a common graded severity scale that is both sensitive to early impairment and to longitudinal changes in patients’ cognitive phenotype, and allows for meaningful comparisons between subgroups of patients, whilst controlling for global severity.

All neuropsychological assessments strongly loaded onto a single unrotated factor, thus providing a single, unified metric of individuals’ global phenotypic severity. This global severity factor demonstrated a good fit across different genetic and diagnostic groups. Furthermore, this severity measure proved to be a more accurate marker of disease progression than the traditional temporal metrics, such as time since diagnosis, testing round, etc. (which proved to be very poor). This data-driven severity measure also allowed temporal metrics to re-enter the analyses as “dependent” variables so as to establish the relative progression “speeds”, within and across patient subgroups, across levels of phenotypic severity.

### Capturing global severity

We were able to validate the measure by examining the distribution of severity scores across at-risk and symptomatic individuals as defined in the GENFI study. As expected, the severity score was higher amongst the symptomatic individuals with a clear gradient reflecting progressive change across the continuum. This measure was also highly stable across the range of tests included, with the same metric emerging from the smaller GENFI 1 or much larger GENFI 2 neuropsychological batteries.

The whole range of severity for symptomatic individuals was also captured across all genetic and diagnostic groups. When comparing to functional impairment, as measured by the CDR®-FTLD score, the severity scores tracked the patients’ increasing functional impairment.^30^ Our analyses indicated that the CDR®-FTLD reflects both cognitive severity as measured by neuropsychological assessments and the reported behavioural changes (CBI-R, FRS).^31^ In the GENFI study, the CDR score is considered when classifying individuals as symptomatic. However, it is useful to examine if a finer gradient of clinical symptoms is present; e.g., mildly symptomatic individuals may overtly resemble the asymptomatic group. Even when behavioural measures are included, our data-driven severity metric captured both global severity and additional variance explained by behavioural measures alone, allowing for more granular severity measurement than CDR®-FTLD.

### Relationship between the severity factor and temporal measures

As age of symptom onset and disease progression varies between and within genetic groups, the clinical spectrum in the GENFI study offered a unique opportunity to investigate the efficacy of disease progression markers. Temporal measures were poorly related to performance on neuropsychological assessments and only explained a very small amount of variance compared to the severity score. When using time since diagnosis as a proxy for disease severity one might expect a direct relationship with worsening symptoms, however we found no correlation.

When monitoring longitudinal change in neurodegenerative disease, measures such as visit number/assessment round are often assumed to act as snapshots of progression. However, as seen in our results, this is not always the case.^30^ When tracked over sequential assessment rounds, cross-sectional samples can be impacted by attrition – both throughout the sample and with biases (e.g., greater attrition of more severe patients). In the GENFI study, like many others, there is a broad range of patients recruited to the study entry, therefore study entry is also not a useful guide to severity. Our results provide further evidence as to why temporal measures are not reliable proxies for disease progression.

### Rates of progression across the severity space

By organising patient data according to the new severity metric, we were then able to revisit time as a dependent measure. This allowed us to examine progression “speed” (time taken to traverse along the severity dimension) and “acceleration”. We observed two main things – one is that the baseline velocity (the intercept in the regression of time onto severity change) across the different genetic types varies, with GRN mutation carriers being the fastest. Secondly, we found that all genetic groups showed gradual and equivalent acceleration across the severity spectrum. This is in line with past studies showing faster decline as cognitive performance worsens and fastest disease progression amongst the GRN genetic carriers.^32–38^ The observed results, consistent with previous research, validate the utility of the phenotypic severity metric to organise patient data and demonstrate how it can enhance our understanding of disease trajectories by reintroducing time as a dependent measure. Consistent with previously reported results, C9orf72 genetic carriers showed the longest time taken to transverse the severity spectrum reflecting previous findings of global impairment, but slower decline.^36,39,40^ The design of clinical trials could be enhanced by the ability to measure an individual’s severity accurately along the continuum. It would also make it possible to schedule follow-up clinical appointments that are either longer or shorter based on anticipated changes in the course of the disease.

### Neuropsychological tests - sensitivity to early clinical changes and a time-efficient sub-battery

The severity score allows a unique way to identify measures sensitive to early clinical changes and in turn potential identification of early decliners. Given that all tests loaded highly onto a single PCA severity dimension, all measures can track severity changes. By examining the spread of scores for each individual test against the global severity metric, we identified the Trail Making Test-B as a potential early marker of cognitive decline. Specifically, a substantial proportion of individuals fell below the normal control-range cut-off for this test, yet had low severity scores. A subset of scores from ‘at-risk’ individuals, who later converted to ‘Symptomatic’, fell into this category. Previous cross-sectional studies have found that cognitively-unimpaired individuals show positive associations between Trail Making Test-B performance and amyloid burden.^41^

The comparison of individual tests to the global severity score also helped to identify specific divergent patterns of performance. Specifically we found that MAPT genetic carriers exhibit early decline on the Boston Naming Test, which is consistent with previously published results of presymptomatic anomia in these individuals.^28,29^

Given that for this cohort, all neuropsychological tests loaded highly on the severity factor, with limited impact of a broader battery, it may not be necessary to run all assessments to estimate severity. Instead there is an opportunity to derive a sub-battery which is very time-efficient yet preserves the ability to capture cognitive severity in this GENFI cohort. Three tests, Digit Symbol, Verbal fluency (letters) and Trail Making Test-B, taking 15min to complete, explained 93% of variance in the cognitive severity score. This type of accurate yet time-efficient sub-battery could help large clinical trials with limited resources and logistical constraints in completing an extensive battery.

### Other data driven approaches

Previous studies have explored interesting data driven methods to develop non-temporal measures of severity. One of these, event-based modelling (EBM) aims to map out decline as a progression of discrete events (e.g., biomarker changes, atrophy patterns) in a serial order using cross-sectional data.^42^ EBM has the underlying assumptions that progression is unidirectional and comprises a series of discrete homogenous events. Accordingly, EBM may be less suitable to modelling neurodegenerative conditions where systems are declining gradually with multiple, overlapping and increasing symptoms which do not form a single, neat serial order with discrete onsets. Discriminative event-based models have been developed to allow for individualised progression sequences, however the events remain independent of each other.^43^ Brain atrophy patterns have also been used to identify subtypes and stages, which model disease processes as fixed, sigmoidal trajectories with probabilistic staging.^9^ This can make it difficult to interpret non-linear rates of change and how they relate to time measures.

### Limitations

The data-driven severity approach explored here does imply several assumptions that should be considered. First, by definition, the resultant PCA metrics (dimension scores) are reflective of the tests included (and excluded), as well as the sample of patients assessed (both the types and the range of severity). Thus, when constructing a model of severity, it is important to assess if severity can be explained by a single or by multidimensional factors. Given the significant augmentation of the neuropsychological test battery between GENFI 1 and 2, we were able to confirm that the single severity factor was unchanged despite the deeper neuropsychological resolution. Interestingly, this same severity factor remained as the first principal component when we added behavioural measures (CBI-R and FRS) to the detailed neuropsychology. In addition, we also observed an emergent second factor which was also a significant predictor of the CDR®-FTLD scores and captured additional variance in behavioural scores. Previous large-scale neuropsychological studies of stroke or neurodegenerative patient groups, using PCA, have shown that dimensions do sometimes vary depending on the patient group and neuropsychological tests included.^44–46^ Future research could look into the challenges associated with dealing with severity as a multidimensional rather than unidimensional construct. One possible approach will be to explore if there are second-order patterns in the deviations away from the first principal severity variation.^18^ This would allow for a systematic way to explore dementia subtypes as it would disentangle which atypical patterns are truly qualitatively different and not simply variations in severity.^18,47–49^

With regard to the spread of patient groups involved, it should be noted that also by definition this data-driven metric of severity pertains to FTD-related genetic-carrying patients. Thus it is a severity metric for this sample alone. Indeed, even within this impressive sample, there was a preponderance of patients with bvFTD. Future explorations of how the method would perform in other aetiologies and more diverse diagnostic groups is warranted to test if meaningful severity comparisons can be made among different neurodegenerative disorders.

## Conclusion

We have shown that a data-driven approach provides an accurate transdiagnostic phenotype-severity metric, which overcomes limitations of traditional temporal markers. It enables mapping individuals along a severity continuum from which one can better identify different disease subtypes in clinical data. This approach has potential applications to interrogate the causes of heterogeneity, and improve clinical trials design: with stratification by severity, and outcome metrics that reveal efficacy of disease-modifying therapy in heterogeneous disorders.

## Supplementary material

Supplementary material is available online.

## Appendix 1

### Consortium

Rhian Convery^1^, Martina Bocchetta^1^, David Cash^1^, Sophie Goldsmith^1^, Kiran Samra^1^, David L. Thomas^2^, Thomas Cope^3^, Maura Malpetti^4^, Antonella Alberici^5^, Enrico Premi^6^, Roberto Gasparotti^7^, Emanuele Buratti^8^, Valentina Cantoni^5^, Andrea Arighi^9^, Chiara Fenoglio^9,10^, Vittoria Borracci^9^, Maria Serpente^9^, Tiziana Carandini^9^, Emanuela Rotondo^9^, Giacomina Rossi^11^, Giorgio Giaccone^11^, Giuseppe Di Fede^11^, Paola Caroppo^11^, Sara Prioni^11^, Veronica Redaelli^11^, David Tang-Wai^12^, Ekaterina Rogaeva^13^, Miguel Castelo-Branco^14^, Morris Freedman^15^, Ron Keren^16^, Sandra Black^17^, Sara Mitchell^17^, Christen Shoesmith^18^, Robart Bartha^19,20^, Rosa Rademakers^21^, Jackie Poos^22^, Janne M. Papma^22^, Lucia Giannini^22^, Liset de Boer^22^, Julie de Houwer^22^, Rick van Minkelen^23^, Yolande Pijnenburg^24^, Benedetta Nacmias^25^, Camilla Ferrari^25^, Cristina Polito^26^, Gemma Lombardi^25^, Valentina Bessi^25^, Enrico Fainardi^27^, Stefano Chiti^27^, Mattias Nilsson^28^, Henrik Viklund^29^, Melissa Taheri Rydell^30,31^, Vesna Jelic^32,33^, Abbe Ullgren^30,31^, Elena Rodriguez-Vieitez^30,31^, Tobias Langheinrich^34,35^, Albert Lladó^36^, Anna Antonell^36^, Jaume Olives^36^, Mircea Balasa^36^, Nuria Bargalló^37^, Sergi Borrego-Ecija^36^, Ana Verdelho^38^, Carolina Maruta^39^, Tiago Costa-Coelho^40-42^, Gabriel Miltenberger^40^, Frederico Simões do Couto^43^, Alazne Gabilondo^44-46^, Ioana Croitoru^45,46^, Mikel Tainta^45,46^, Myriam Barandiaran^44-46^, Patricia Alves^45,47^, Benjamin Bender^48^, David Mengel^49,50^, Lisa Graf^49^, Annick Vogels^51^, Mathieu Vandenbulcke^52^, Philip Van Damme^53,54^, Rose Bruffaerts^55,56^, Koen Poesen^57^, Pedro Rosa-Neto^58^, Maxime Montembault^59^, Raffaella Lara Migliaccio^60,61^, Ninon Burgos^60,62^, Daisy Rinaldi^60,61^, Catharina Prix^63^, Elisabeth Wlasich^63^, Olivia Wagemann^63^, Sonja Schönecker^63^, Alexander Maximilian Bernhardt^63^, Anna Stockbauer^63^, Jolina Lombardi^64^, Sarah Anderl-Straub^64^, Adeline Rollin^65^, Gregory Kuchcinski^66^, Vincent Deramecourt^66^, João Durães^67^, Marisa Lima^67^, Maria João Leitão^68^, Maria Rosario Almeida^69^, Miguel Tábuas-Pereira^67,69^, Sónia Afonso^70^, João Lemos^69^, Erdi Şahin^71^, Sanem Sultan Yoruk Oner^71^, Duygu Aybar^71^

### Affiliations

^1^Department of Neurodegenerative Disease, Dementia Research Centre, UCL Queen Square Institute of Neurology, London, UK

^2^Neuroimaging Analysis Centre, Department of Brain Repair and Rehabilitation, UCL Institute of Neurology, Queen Square, London, UK

^3^Cambridge University Hospitals NHS Trust, Cambridge UK

^4^Department of Clinical Neurosciences, University of Cambridge, Cambridge, UK

^5^Centre for Neurodegenerative Disorders, Department of Clinical and Experimental Sciences, University of Brescia, Brescia, Italy

^6^Stroke Unit, ASST Brescia Hospital, Brescia, Italy

^7^Neuroradiology Unit, University of Brescia, Brescia, Italy

^8^ICGEB Trieste, Italy

^9^Fondazione IRCCS Ca’ Granda Ospedale Maggiore Policlinico, Neurodegenerative Diseases Unit, Milan, Italy

^10^University of Milan, Centro Dino Ferrari, Milan, Italy

^11^Fondazione IRCCS Istituto Neurologico Carlo Besta, Milano, Italy

^12^The University Health Network, Krembil Research Institute, Toronto, Canada

^13^Tanz Centre for Research in Neurodegenerative Diseases, University of Toronto, Toronto, Canada

^14^Faculty of Medicine, ICNAS, CIBIT, University of Coimbra, Coimbra, Portugal

^15^Baycrest Health Sciences, Rotman Research Institute, University of Toronto, Toronto, Canada

^16^The University Health Network, Toronto Rehabilitation Institute, Toronto, Canada

^17^Sunnybrook Health Sciences Centre, Sunnybrook Research Institute, University of Toronto, Toronto, Canada

^18^Department of Clinical Neurological Sciences, University of Western Ontario, London, Ontario, Canada

^19^Department of Medical Biophysics, The University of Western Ontario, London, Ontario, Canada

^20^Centre for Functional and Metabolic Mapping, Robarts Research Institute, The University of Western Ontario, London, Ontario, Canada

^21^Center for Molecular Neurology, University of Antwerp, Antwerp, Belgium

^22^Department of Neurology, Erasmus Medical Center, Rotterdam, Netherlands

^23^Department of Clinical Genetics, Erasmus Medical Center, Rotterdam, Netherlands

^24^Amsterdam University Medical Centre, Amsterdam VUMC, Amsterdam, Netherlands

^25^Department of Neuroscience, Psychology, Drug Research and Child Health, University of Florence, Florence, Italy

^26^Department of Biomedical, Experimental and Clinical Sciences “Mario Serio”, Nuclear Medicine Unit, University of Florence, Florence, Italy

^27^Neuroradiology Unit, Department of Experimental and Clinical Biomedical Sciences, University of Florence, Florence, Italy

^28^Department of Clinical Neuroscience, Karolinska Institutet, Stockholm, Sweden

^29^Karolinska University Hospital Huddinge, Sweden

^30^Department of Neurobiology, Care Sciences and Society; Center for Alzheimer Research, Division of Neurogeriatrics, Bioclinicum, Karolinska Institutet, Solna, Sweden

^31^Unit for Hereditary Dementias, Theme inflammation and Aging, Karolinska University Hospital, Solna, Sweden

^32^Department of Neurobiology, Care Sciences and Society; Division of Clinical Geriatrics, Karolinska Institutet, Stockholm, Sweden

^33^Cognitive clinic, Theme inflammation and Aging, Karolinska University Hospital, Solna, Sweden

^34^Division of Neuroscience and Experimental Psychology, Wolfson Molecular Imaging Centre, University of Manchester, Manchester, UK

^35^Manchester Centre for Clinical Neurosciences, Department of Neurology, Salford Royal NHS Foundation Trust, Manchester, UK

^36^Alzheimer’s disease and Other Cognitive Disorders Unit, Neurology Service, Hospital Clínic, Barcelona, Spain

^37^Imaging Diagnostic Center, Hospital Clínic, Barcelona, Spain

^38^Department of Neurosciences and Mental Health, Centro Hospitalar Lisboa Norte - Hospital de Santa Maria & Faculty of Medicine, University of Lisbon, Lisbon, Portugal

^39^Laboratory of Language Research, Centro de Estudos Egas Moniz, Faculty of Medicine, University of Lisbon, Lisbon, Portugal

^40^Faculty of Medicine, University of Lisbon, Lisbon, Portugal

^41^Institute of Molecular Medicine João Lobo Antunes, University of Lisbon, Lisbon, Portugal

^42^Research Institute for Medicines, Faculty of Pharmacy, University of Lisbon, Lisbon, Portugal

^43^Faculdade de Medicine, Universidade Catolica Portuguesa, Rio de Mouro, Portugal

^44^Cognitive Disorders Unit, Department of Neurology, Donostia University Hospital, San Sebastian, Gipuzkoa, Spain

^45^Instituto de Investigación Sanitaria Biogipuzkoa, Neurosciences Area, Group of Neurodegenerative Diseases, San Sebastian, Spain

^46^Center for Biomedical Research in Neurodegenerative Disease (CIBERNED), Carlos III Health Institute, Madrid, Spain

^47^Department of Educational Psychology and Psychobiology, Faculty of Education, International University of La Rioja, Logroño, Spain

^48^Department of Diagnostic and Interventional Neuroradiology, University of Tübingen, Tübingen, Germany

^49^Department of Neurodegenerative Diseases, Hertie-Institute for Clinical Brain Research and Center of Neurology, University of Tübingen, Tübingen, Germany

^50^Center for Neurodegenerative Diseases (DZNE), Tübingen, Germany

^51^Department of Human Genetics, KU Leuven, Leuven, Belgium

^52^Geriatric Psychiatry Service, University Hospitals Leuven, Belgium; Neuropsychiatry, Department of Neurosciences, KU Leuven, Leuven, Belgium

^53^Neurology Service, University Hospitals Leuven, Belgium

^54^Laboratory for Neurobiology, VIB-KU Leuven Centre for Brain Research, Leuven, Belgium

^55^Department of Biomedical Sciences, University of Antwerp, Antwerp, Belgium

^56^Biomedical Research Institute, Hasselt University, Hasselt, Belgium

^57^Laboratory for Molecular Neurobiomarker Research, KU Leuven, Leuven, Belgium

^58^Translational Neuroimaging Laboratory, McGill Centre for Studies in Aging, McGill University, Montreal, Québec, Canada

^59^Douglas Research Centre, Department of Psychiatry, McGill University, Montreal, Québec, Canada

^60^Sorbonne Université, Paris Brain Institute – Institut du Cerveau – ICM, Inserm U1127, CNRS UMR 7225, AP-HP - Hôpital Pitié-Salpêtrière, Paris, France

^61^Centre de référence des démences rares ou précoces, IM2A, Département de Neurologie, AP-HP - Hôpital Pitié-Salpêtrière, Paris, France

^62^Inria, Aramis project-team, F-75013, Paris, France; Centre pour l’Acquisition et le Traitement des Images, Institut du Cerveau et la Moelle, Paris, France

^63^Neurologische Klinik, Ludwig-Maximilians-Universität München, Munich, Germany

^64^Department of Neurology, University of Ulm, Ulm, Germany

^65^CHU, CNR-MAJ, Labex Distalz, LiCEND Lille, France

^66^Lille Neuroscience & Cognition U1172, University of Lille, Inserm, CHU Lille, France

^67^Neurology Department, Centro Hospitalar e Universitario de Coimbra, Coimbra, Portugal

^68^Centre of Neurosciences and Cell Biology, Universidade de Coimbra, Coimbra, Portugal

^69^Faculty of Medicine, University of Coimbra, Coimbra, Portugal

^70^Instituto Ciencias Nucleares Aplicadas a Saude, Universidade de Coimbra, Coimbra, Portugal

^71^Behavioral Neurology and Movement Disorders Unit, Istanbul Faculty of Medicine, Istanbul University, Istanbul, Turkiye

