## Supplementary Tables for "Time’s up: Using data-driven phenotype-severity metrics not time to map progression in the dementias"

**Supplementary Table 1: Descriptive table of neuropsychological assessments and missing data across all visits for symptomatic and converter individuals pre-imputation.**

| Visit number | 1  (*n* = 265) | 2  (*n* = 132) | 3  (*n* = 61) | 4  (*n* = 27) | 5  (*n* = 17) | 6  (*n* = 13) | 7  (*n* = 4) | 8  (*n* = 3) |
| --- | --- | --- | --- | --- | --- | --- | --- | --- |
| MMSE,  mean (SD)  median [Min, Max]  missing n (%) | 23.2 (6.4)  25.0 [1.0, 30.0]  17 (6.4%) | 22.3 (7.2)  25.0 [2.0, 30.0]  11 (8.3%) | 24.0 (6.2)  26.0 [2.0, 30.0]  4 (6.6%) | 24.2 (5.2)  25.0 [11.0, 30.0]  3 (11.1%) | 25.3 (4.89)  27.5 [13.0, 30.0]  1 (5.9%) | 24.3 (5.12)  25.0 [13.0, 30.0]  0 (0%) | 27.0 (2.2)  26.5 [25.0, 30.0]  0 (0%) | 25.7 (3.1)  25.0 [23.0, 29.0]  0 (0%) |
| Digit Span Forward,  mean (SD)  median [Min, Max]  missing n (%) | 5.5 (2.5)  6.0 [0, 12.0]  3 (1.1%) | 5.7 (2.8)  6.0 [0, 12.0]  1 (0.8%) | 6.0 (2.5)  6.0 [2.0, 12.0]  5 (8.2%) | 6.2 (2.4)  6.0 [1.0, 12.0]  0 (0%) | 6.5 (2.0)  6.0 [4.0, 11.0]  2 (11.8%) | 6.2 (2.1)  6.0 [4.0, 10.0]  2 (15.4%) | 7.5 (3.1)  7.5 [4.0, 11.0]  0 (0%) | 6.7 (4.2)  8.0 [2.0, 10.0]  0 (0%) |
| Digit Span Backward,  mean (SD)  median [Min, Max]  missing n (%) | 3.7 (2.3)  4.0 [0, 11.0]  3 (1.1%) | 4.0 (2.4)  4.0 [0, 11.0]  5 (3.8%) | 4.0 (2.3)  4.0 [0, 10.0]  5 (8.2%) | 4.4 (2.1)  5.0 [0, 8.0]  0 (0%) | 4.7 (1.6)  5.0 [3.0, 9.0]  2 (11.8%) | 4.7 (2.4)  4.0 [2.0, 10.0]  2 (15.4%) | 6.3 (3.3)  6.0 [3.0, 10.0]  0 (0%) | 6.7 (3.5)  7.0 [3.0, 10.0]  0 (0%) |
| Digit Symbol,  mean (SD)  median [Min, Max]  missing n (%) | 27.0 (15.0)  26.5 [0, 68.0]  15 (5.7%) | 30.4 (17.1)  29.0 [0, 93.0]  19 (14.4%) | 32.6 (17.0)  28.5 [0, 79.0]  9 (14.8%) | 32.9 (16.6)  29.0 [9.0, 75.0]  3 (11.1%) | 30.9 (18.3)  28.5 [8.0, 80.0]  3 (17.6%) | 29.5 (20.2)  28.0 [7.0, 80.0]  3 (23.1%) | 28.3 (12.2)  31.5 [11.0, 39.0]  0 (0%) | 34.3 (8.1)  33.0 [27.0,43.0]  0 (0%) |
| Boston Naming,  mean (SD)  median [Min, Max]  missing n (%) | 20.3 (7.9)  23.0 [0, 30.0]  9 (3.4%) | 19.6 (8.7)  21.5 [0, 30.0]  3 (2.3%) | 20.8 (7.9)  23.0 [0, 30.0]  0 (0%) | 21.8 (7.1)  24.0 [7.0, 30.0]  1 (3.7%) | 22.5 (7.2)  24.0 [5.0, 30.0]  0 (0%) | 20.2 (8.6)  22.0 [4.0, 30.0]  0 (0%) | 23.0 (5.6)  22.0 [18.0, 30.0]  0 (0%) | 20.7 (8.5)  21.0 [12.0, 29.0]  0 (0%) |
| Block Design,  mean (SD)  median [Min, Max]  missing n (%) | 19.7 (16.5)  16.0 [0, 67.0]  35 (13.2%) | 21.6 (18.4)  20.0 [0, 62.0]  33 (25%) | 24.0 (19.7)  17.5 [0, 60.0]  13 (21.3%) | 26.2 (20.3)  22.5 [0, 57.0]  5 (18.5%) | 28.5 (19.8)  23.0 [0, 61.0]  3 (17.6.1%) | 24.0 (21.8)  15.0 [2.0, 59.0]  4 (30.8%) | 26.5 (20.2)  29.5 [2.0, 45.0]  0 (0%) | 41.5 (6.4)  41.5 [37.0, 46.0]  1 (33.3%) |
| Verbal Fluency - Animals,  mean (SD)  median [Min, Max]  missing n (%) | 11.9 (7.04)  11.0 [0, 39.0]  10 (3.8%) | 12.0 (7.42)  11.0 [0, 34.0]  7 (5.3%) | 13.5 (7.6)  13.0 [0, 28.0]  1 (1.6%) | 13.4 (7.4)  13.0 [0, 27.0]  2 (7.4%) | 12.4 (5.9)  11.0 [5.0, 24.0]  1 (5.9%) | 10.3 (7.0)  7.0 [1.0, 20.0]  0 (0%) | 11.5 (5.8)  10.5 [6.0, 19.0]  0 (0%) | 12.3 (5.5)  15.0 [6.0, 16.0]  0 (0%) |
| Verbal Fluency - Letters,  mean (SD)  median [Min, Max]  missing n (%) | 18.5 (14.0)  16.0 [0, 77.0]  14 (5.3%) | 18.5 (14.2)  17.0 [0, 63.0]  1 (8.3%) | 23.0 (15.6)  20.0 [0, 66.0]  4 (6.6%) | 28.8 (17.7)  23.0 [6.00, 64.0]  3 (11.1%) | 25.5 (14.4)  21.0 [8.0, 50.0]  2 (11.8%) | 24.3 (15.9)  21.5 [7.0, 54.0]  1 (7.7%) | 29.0 (14.8)  28.0 [12.0, 48.0]  0 (0%) | 31.7 (13.2)  29.0 [20.0, 46.0]  0 (0%) |
| Trail Making – A time,  mean (SD)  median [Min, Max]  missing n (%) | 66.8 (41.9)  50.0 [150, 14.0)  14 (5.3%) | 64.2 (37.3)  58.0 [150, 15.0]  15 (11.4%) | 60.2 (35.6)  52.0 [150, 17.0]  2 (3.3%) | 65.6 (39.1)  52.0 [150, 15.0]  0 (0%) | 57.9 (39.2)  41.0 [150, 14.0]  1 (5.9%) | 65.5 (39.2)  58.5 [150, 14.0]  1 (7.7%) | 49.3 (26.2)  40.0 [87.0, 30.0]  0 (0%) | 43.7 (15.5)  39.0 [61.0, 31.0]  0 (0%) |
| Trail Making – B time,  mean (SD)  median [Min, Max]  missing n (%) | 198 (94.7)  203 [300, 34]  41 (15.5%) | 189 (96.8)  180 [300, 10]  29 (22%) | 169 (94.1)  135 [300, 46)  8 (13.1%) | 176 (96.7)  176 (300, 43)  5 (18.5%) | 166 (102)  126 [300, 38]  2 (11.8%) | 136 (104)  90.5 [300, 39]  5 (38.5%) | 132 (60.0)  103 [201, 92]  1 (25.0%) | 141 (62.3)  153 [197, 74]  0 (0%) |

| A) All visits | PC1 | PC2 | PC3 | PC4 | PC5 |
| --- | --- | --- | --- | --- | --- |
| Block Design | 0.857 |  | -0.255 |  | 0.265 |
| Digit Symbol | 0.900 |  | -0.212 |  |  |
| Digit Span Forward | 0.638 | 0.570 | 0.440 |  | 0.212 |
| Digit Span Backward | 0.785 | 0.419 |  |  | -0.372 |
| Trail Making A-time | 0.822 |  | -0.176 | 0.434 | -0.132 |
| Trail Making B-time | 0.880 |  | -0.179 |  | 0.145 |
| Boston Naming | 0.627 | -0.534 | 0.505 | 0.174 |  |
| Verbal Fluency-Animals | 0.840 | -0.335 |  | -0.242 |  |
| Verbal Fluency-Letters | 0.853 |  |  | -0.379 |  |
| Proportion variance | 65% | 10% | 7% | 5% | 3% |
| Eigenvalue | 5.85 | 0.92 | 0.63 | 0.44 | 0.30 |

**Supplementary Table 2:** **Principal Component Analyses (PCA) results for the smaller core neuropsychological battery across GENFI 1 and GENFI 2. A)** including all visits for symptomatic and converter individuals; **B)** including first baseline visits only; **C)** including first, middle and last visits only. Columns display neuropsychological assessment loadings for each component, with the bottom two rows reflecting eigenvalues and variance explained.

| B) First visit only | PC1 | PC2 | PC3 | PC4 | PC5 |
| --- | --- | --- | --- | --- | --- |
| Block Design | 0.821 | -0.135 | -0.316 |  | 0.378 |
| Digit Symbol | 0.894 |  | -0.223 |  |  |
| Digit Span Forward | 0.645 | -0.524 | 0.488 | 0.111 | 0.108 |
| Digit Span Backward | 0.811 | -0.362 | 0.109 |  | -0.172 |
| Trail Making A-time | 0.820 |  | -0.166 | 0.387 | -0.313 |
| Trail Making B-time | 0.867 |  | -0.189 |  |  |
| Boston Naming | 0.626 | 0.590 | 0.406 | 0.216 | 0.139 |
| Verbal Fluency-Animals | 0.834 | 0.338 |  | -0.274 |  |
| Verbal Fluency-Letters | 0.846 |  |  | -0.394 | -0.135 |
| Proportion variance | 64% | 10% | 7% | 5% | 4% |
| Eigenvalue | 5.78 | 0.90 | 0.64 | 0.45 | 0.32 |

| C) First, middle and last visits | PC1 | PC2 | PC3 | PC4 | PC5 |
| --- | --- | --- | --- | --- | --- |
| Block Design | 0.877 |  | -0.128 |  | -0.166 |
| Digit Symbol | 0.908 | -0.113 | -0.140 | -0.179 |  |
| Digit Span Forward | 0.563 | 0.655 | 0.343 | 0.276 | -0.210 |
| Digit Span Backward | 0.700 | 0.474 |  | -0.113 | 0.508 |
| Trail Making A-time | 0.833 |  |  | -0.383 |  |
| Trail Making B-time | 0.882 |  |  | -0.164 | -0.218 |
| Boston Naming | 0.626 | -0.547 | 0.509 |  | 0.131 |
| Verbal Fluency-Animals | 0.818 | -0.349 |  | 0.269 |  |
| Verbal Fluency-Letters | 0.818 |  | -0.301 | 0.345 | 0.108 |
| Proportion variance | 62% | 12% | 6% | 5% | 5% |
| Eigenvalue | 5.60 | 1.10 | 0.52 | 0.49 | 0.41 |

**Supplementary Table 3: Results from the McNemar test comparing two types of discrepancies between test scores and severity ratings.** 1) test score above the impaired threshold but severity score indicates high severity (false positives); 2) test score below the impaired threshold but severity score indicates low severity (false negatives). True positives are cases where the test score is above the impaired threshold and the severity score indicates low severity, while true negatives are cases where the test score is below the impaired threshold and the severity score indicates high severity.

| Test | True positive | False positive | False negative | True negative | McNemar statistic | p-value |
| --- | --- | --- | --- | --- | --- | --- |
| Boston Naming | 1199 | 11 | 261 | 207 | 227.95 | *P* < 0.001 |
| Trail Making B-time | 1340 | 3 | 120 | 215 | 109.40 | *P* < 0.001 |
| Trail Making A-time | 1415 | 46 | 45 | 172 | 0 | *P* = 1 |
| Block Design | 1420 | 58 | 40 | 160 | 2.95 | *P* = 0.09 |
| Digit Symbol | 1456 | 114 | 4 | 104 | 100.69 | *P* < 0.001 |
| Digit Span Forward | 1436 | 94 | 24 | 124 | 40.35 | *P* < 0.001 |
| Digit Span Backward | 1427 | 40 | 33 | 178 | 0.49 | *P* = 0.48 |
| Verbal Fluency-Animals | 1408 | 44 | 52 | 174 | 0.51 | *P* = 0.52 |
| Verbal Fluency-Letters | 1381 | 23 | 79 | 195 | 29.66 | *P* < 0.001 |

**Supplementary Table 4: Results of the Principal Component Analysis (PCA) for the extended GENFI 2 neuropsychological battery across symptomatic and converter individuals.** Columns display neuropsychological assessment loadings for each component, with the bottom two rows reflecting eigenvalues and variance explained.

| GENFI 2 | PC1 | PC2 | PC3 | PC4 | PC5 |
| --- | --- | --- | --- | --- | --- |
| Block Design | 0.794 | -0.237 | 0.109 | -0.210 | 0.138 |
| Digit Symbol | 0.852 |  |  | -0.189 |  |
| Digit Span Forward | 0.482 | -0.507 | 0.293 |  | 0.177 |
| Digit Span Backward | 0.668 | -0.428 | 0.248 |  |  |
| Trail Making A-time | 0.775 | -0.212 |  |  |  |
| Trail Making B-time | 0.836 | -0.202 |  | -0.209 |  |
| Boston Naming | 0.667 | 0.377 | -0.387 |  |  |
| Verbal Fluency-Animals | 0.831 | 0.180 |  | -0.146 | -0.142 |
| Verbal Fluency-Letters | 0.811 |  | 0.229 | -0.105 | -0.162 |
| Camel and Cactus | 0.811 | 0.228 | -0.256 | 0.103 |  |
| Benson figure copy | 0.586 | -0.201 |  | 0.382 | 0.464 |
| Benson figure recall | 0.587 | 0.115 | -0.496 | -0.203 | 0.459 |
| Free and Cued Selective Reminding Test | 0.800 | 0.183 | -0.253 | -0.258 |  |
| Stroop colour-total | 0.607 | -0.135 | -0.228 | 0.541 | -0.188 |
| Stroop colour-time | 0.806 |  | 0.114 |  | -0.298 |
| Stroop ink-total | 0.700 | -0.237 |  | 0.427 |  |
| Stroop ink-time | 0.787 |  | 0.113 | -0.216 | -0.238 |
| Faux pas | 0.806 |  |  | 0.126 | -0.186 |
| Ekman | 0.839 |  |  |  |  |
| Modified Interpersonal Reactivity Index | 0.416 | 0.588 | 0.504 |  | 0.287 |
| Revised Self-Monitoring Scale | 0.537 | 0.604 | 0.351 | 0.181 | 0.135 |
| Proportion variance | 53% | 8% | 6% | 5% | 4% |
| Eigenvalue | 11.06 | 1.71 | 1.22 | 1.02 | 0.87 |

**Supplementary Table 5: Results of the PCA for the core neuropsychological battery (GENFI 1 and GENFI 2) additionally including CBI and FRS measures across symptomatic and converter individuals**. Columns display neuropsychological assessment loadings for each component, with the bottom two rows reflecting eigenvalues and variance explained.

| GENFI 1 + GENFI 2 | PC1 | PC2 | PC3 | PC4 | PC5 |
| --- | --- | --- | --- | --- | --- |
| Block Design | 0.848 | -0.124 | 0.136 | -0.219 |  |
| Digit Symbol | 0.900 |  |  | -0.233 |  |
| Digit Span Forward | 0.587 | -0.524 | 0.201 | 0.505 | 0.133 |
| Digit Span Backward | 0.746 | -0.404 | 0.230 | 0.158 |  |
| Trail Making A-time | 0.813 | -0.109 |  | -0.258 | 0.405 |
| Trail Making B-time | 0.862 | -0.174 |  | -0.211 |  |
| Boston Naming | 0.627 |  | -0.708 | 0.223 | 0.132 |
| Verbal Fluency-Animals | 0.862 | 0.154 | -0.226 |  | -0.232 |
| Verbal Fluency-Letters | 0.854 |  |  |  | -0.373 |
| Cambridge Behavioural Inventory  Frontotemporal Dementia Rating Scale | 0.571  0.595 | 0.736  0.702 | 0.155  0.271 | 0.171  0.117 | 0.101 |
| Proportion variance | 58% | 14% | 7% | 5% | 4% |
| Eigenvalue | 6.38 | 1.56 | 0.77 | 0.59 | 0.42 |
