## Supplementary Figures for "Time’s up: Using data-driven phenotype-severity metrics not time to map progression in the dementias"

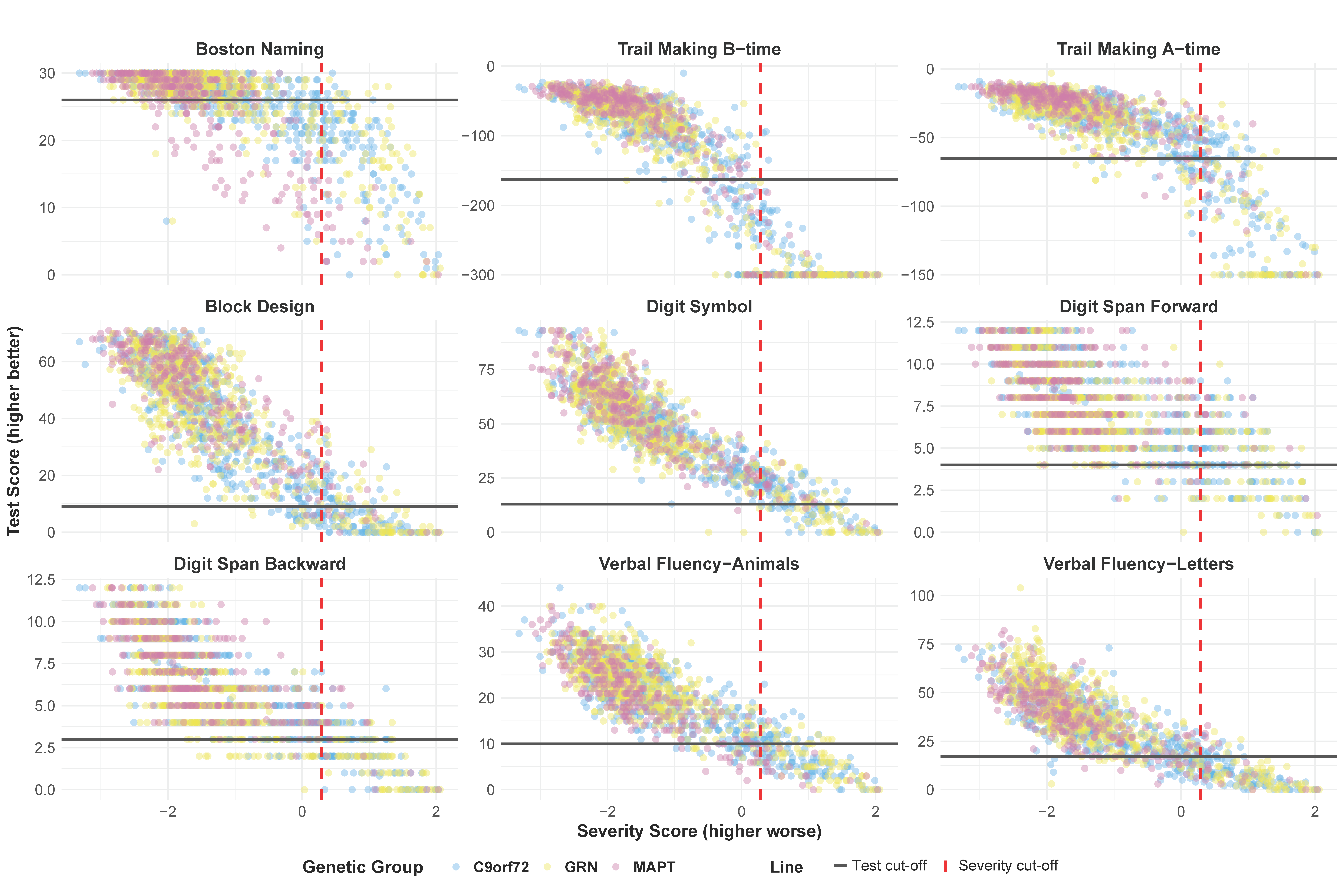
**Supplementary Figure 1:** **Association of the overall severity score with raw scores of neuropsychological tests and their cut-offs stratified by genetic group.** A lower severity score indicates less impairment. Black horizontal lines represent the test score cut-off selected at 1.5 SD below the mean for a 72 year old with 12 years of education. The vertical red dashed line is the severity cut-off calculated by all test score cut-off’s multiplied by test weightings on the first component.

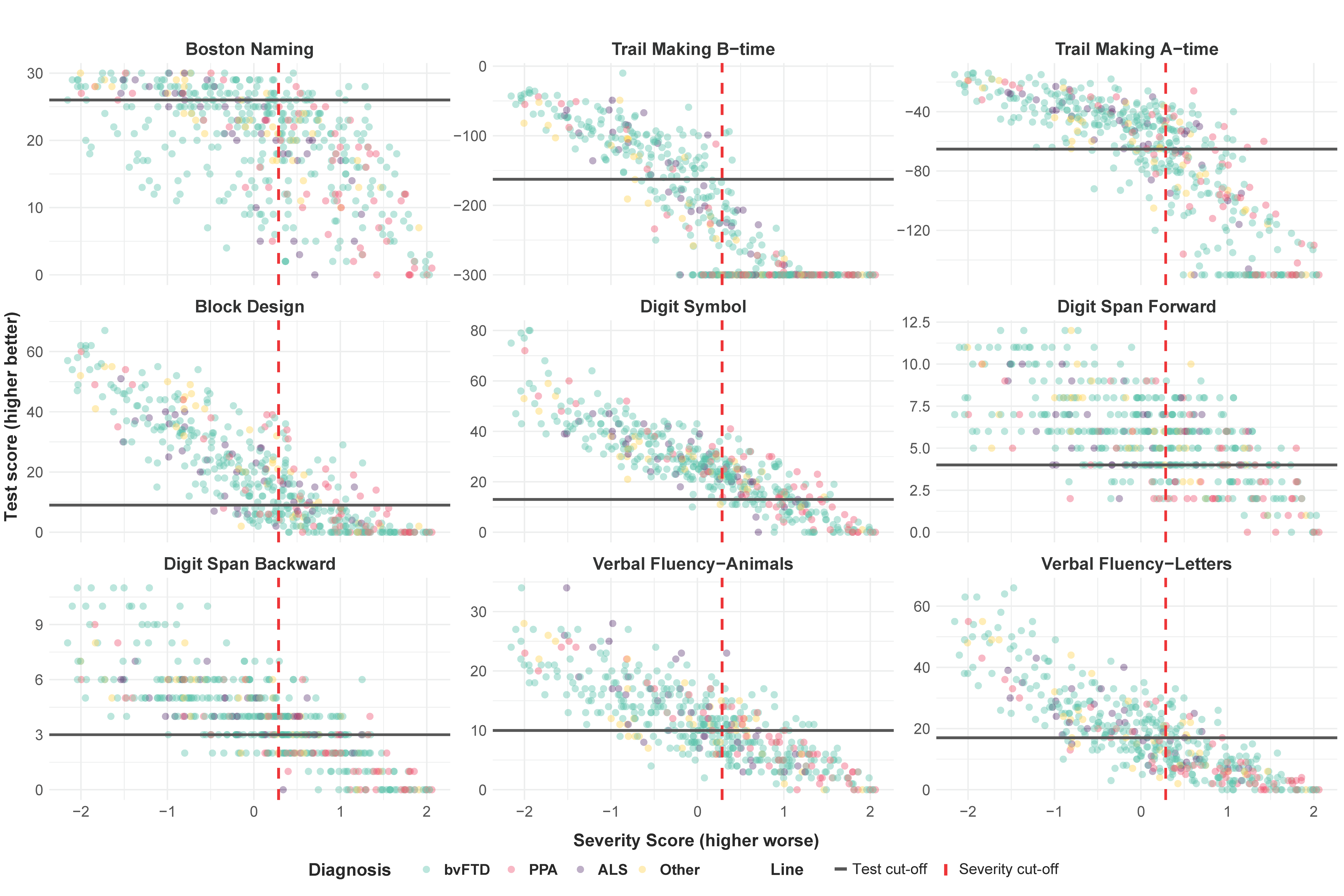
**Supplementary Figure 2: Association of the overall severity score with raw scores of neuropsychological tests and their cut-offs stratified by diagnostic group**. A lower severity score indicates less impairment. Black horizontal lines represent the test score cut-off selected at 1.5 SD below the mean for a 72 year old with 12 years of education. The vertical red dashed line is the severity cut-off calculated by all test score cut-off’s multiplied by test weightings on the first component.

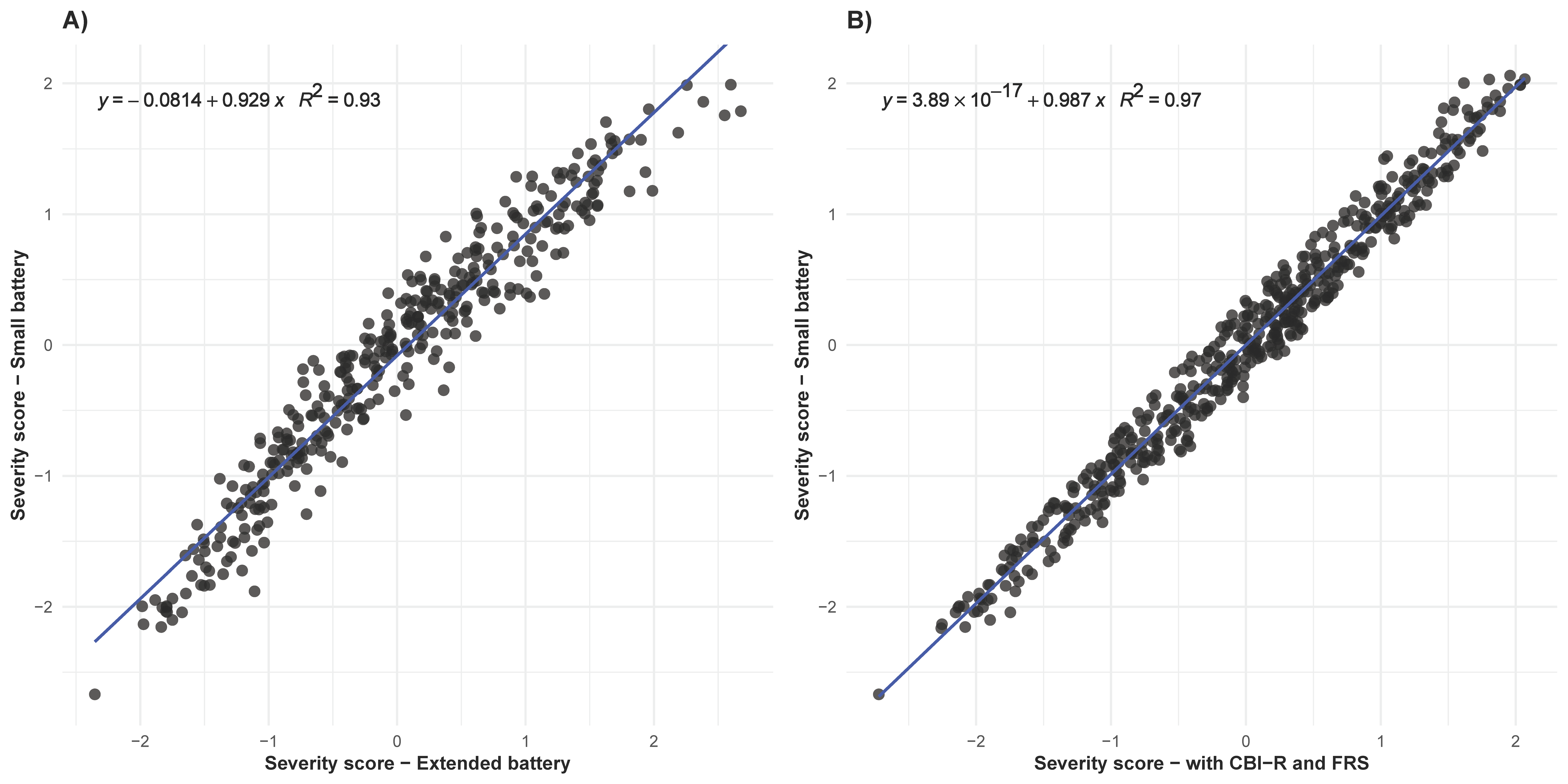

**Supplementary Figure 3:** **Severity score comparisons across extended assessment batteries.** Severity scores for symptomatic individuals were compared A) to using the small assessment battery (GENFI 1) and a larger assessment battery (GENFI 2); B) to using the small assessment battery (GENFI 1) and when additionally including the Cambridge Behavioural Inventory-Revised (CBI-R) and the Frontotemporal Dementia Rating Scale (FRS). Every point represents a participant. Each fitted linear regression equation and *R²* value show a strong within-individual correlation between severity scores from the two assessment batteries.
